# Efficacy and Safety of Indomethacin in Covid-19 patients

**DOI:** 10.1101/2020.12.14.20245266

**Authors:** Rajan Ravichandran, Prassana Purna, Sivakumar Vijayaragavan, Ravi Teja Kalavakollu, Shilpa Gaidhane, Ramarathnam Krishna Kumar

**Author notes:** **Conflict of interest** The authors have no conflicts of interest to report. **Funding:** Funding for this study was provided by Mr. Kris Gopalakrishnan, Alumnus of the Indian Institute of Technology Madras. He is also the Chairman of Axilor Ventures and former executive vice chairman (former co-chairman) of Infosys.

## Abstract

**Background:** Indomethacin, a well-known non-steroidal anti-inflammatory drug (NSAID), has in addition broad spectrum anti-viral activity in the laboratory including on SARS-Cov-2 virus. This trial is to observe the likelihood of efficacy and safety of Indomethacin in treating RT-PCR positive Covid patients

**Materials and Methods:** The study was done in two groups. In the first group of the study, where mild and moderate patients were involved, Propensity Score Matching was used as a methodology to compare Indomethacin and paracetamol based treatments in addition to the standard protocol for treating covid-19 patients.. Blood chemistry was collected before and after the treatment. The patients were monitored every day for clinical parameters. In this part, a patient developing hypoxia was the end point. In the second group, severe patients admitted with hypoxia were treated with Indomethacin in addition to Remdesivir, and the end point was the requirement of mechanical ventilation/admission to ICU.

**Results:** It was observed that patients treated with Indomethacin had a reduction in the number of days to become afebrile, reduction in cough and myalgia by half compared to the paracetamol set. Only one out of 72 patients in the Indomethacin arm of the first group required supplementary oxygen while 28 of the 72 patients required supplementary oxygen in the paracetamol arm. No one in the second group deteriorated enough to require mechanical ventilation. There wea no evidence of adverse reaction to indomethacin or deterioration of renal or liver function.

**Conclusion:** Indomethacin, along with standard care, seems to provide faster symptomatic relief and prevent progression of pneumonia in Covid-19 patients. It should be considered to replace paracetamol when there is no contraindication for its use.

## 1.0 Introduction

SARS–Cov-2, or Covid-19, which belongs to the family of coronavirus, has led to a pandemic the likes of which the world has not seen in a hundred years. Though the mechanism of the virus-host interaction and the possible treatment have been the subject of several hundred studies, effective and safe treatment regime or a vaccine is yet to emerge. Drug repurposing may be a possible solution for immediate treatment, and scores of drugs have been suggested from various perspectives [1,2]

The drugs that could be used for the treatment of Covid-19 can be classified into three categories: Antivirals, anti-inflammatory and supporting therapies. Antiviral action can further be broadly classified into three categories [3]. The first category of anti-virals is the fusion inhibitors, or virus-entry preventers. These essentially prevent the fusion of the virus and the penetration into the human cell cytoplasm [3]. Enzymes such as the Angiotensin-Converting Enzyme (ACE-2) and the Transmembrane serine protease (TMPRSS2), play important roles in endocytosis. One of the key factors in virus entry is the protease activation of S glycoprotein. Cathepsin L activity is crucial in this step and inhibiting Cathepsin L inhibits the entry of SARS-Cov-2 by 76 % [4]. The drugs of choice as fusion-inhibitors so far are Baricitinib, Camostat mesylate and Umifenovir. But none has shown the required efficacy for treatment.

Proteases such as M^pro^, P^pro^ and 3C-like cysteine proteases play a central role in the replication and transcription processes. During replication and particle assembly, viral proteases cleave polyproteins expressed by the virus to produce a number of essential Non-Structural Proteins (NSPs). Hence, protease-inhibitors are a popular class of antiviral candidates [3]. The end result of viral transcription is the production of a negative sense RNA that is used for replication. The formation of RNA-dependent RNA polymerase (RdRp), a crucial enzyme in the replication process, is another target for drug discovery. Two sets of drugs have been tried to blunt this step. Ritonavir-Lopinavir combination, for example, works to block the formation of NSPs. They are protease inhibitors. Another set of drugs, such as Remdesivir and Favipiravir, interferes with the RdRp thus, inhibiting virus replication.

Pro-inflammatory cytokine production is a natural process during an immune response. An important step in the control of the disease is the elimination of virus-infected cells. If this step, which naturally follows virus entry and replication, is defective or prolonged, several pro-inflammatory cytokines are generated in an uncontrolled fashion resulting in what is popularly called a “cytokine storm” [5]. Several Interleukins are involved in a “cytokine storm”, the foremost among them are IL-6, IL-1 [5] and IL-17 [6]. IL-17 also seems to have a role as the interaction partner of SARS-Cov-2. Anti-inflammatory drugs targeting the production of these Interleukins are an important choice for treatment.

### 1.1 Indomethacin as a drug for SARS-Cov-2

The first paper that showed the anti-viral property of Indomethacin was in 2006 [7]. The following is an account of the role of Indomethacin in the pathogenic cycle. The viral entry can be inhibited by down regulating ACE2 and TMPRSS2 [8]. Using an open source code, Gene2Drug, Napolitano *et al*., [8] found that Indomethacin indeed down regulates ACE2 by suppressing the genes in the ACE2 pathway. The other factor, as stated earlier, is the inhibition of Cathepsin L activity for fusion. Ragav *et al*. [9] showed that Indomethacin inhibits Cathepsin L activity and no other non-steroidal anti-inflammatory drug has any role in the Cathepsin L activity. Hence, theoretically, Indomethacin could be a major candidate as a fusion inhibitor. The role of Nsp7, a cofactor of Nsp12 for RNA synthesis, has been highlighted by Frediansyah *et al*. [2]. Gordan *et al*. [6] identified PGES-2 as the “interactor” with Nsp7 and Indomethacin being a Prostaglandin E synthase 2 (PGES-2) inhibitor, is a candidate for blocking RNA synthesis. That it blocks RNA synthesis was also shown by Amici *et al*. [7].

The anti-inflammatory effect of Indomethacin is well understood. IL-6, a key Interleukin, and its surrogate C-Reactive Protein (CRP), are raised in Covid-19 patients. Indomethacin is known to reduce IL-6 by inhibiting the synthesis of PGES-2 [10]. The role of Indomethacin in lowering IL-6 in SARS-CoV-2 patients has been highlighted by Russel *et al*. [11]. Indomethacin has been used successfully to prevent cytokine reaction in kidney transplant patients receiving OKT3 therapy [12,13].

There is experimental evidence of the effectiveness of Indomethacin *in vitro* against SARS-CoV-1 by Amici *et al*. [6]. Direct evidence for SARS-Cov-2 is provided by Xu *et al*. [14]. They have shown the antiviral effect of Indomethacin *in vitro, in cellulo* and in Corona-infected canine model, though they state that Indomethacin does not reduce infectivity, binding or entry into target cells. Such a conclusion comes from an earlier study [7] and needs to be verified as the mechanisms suggested for blocking virus fusion are very robust.

Though there have been suggestions in many of the above-mentioned publications, no proper clinical trial to evaluate Indomethacin has been carried out. In a very recent paper, Gordon *et al* [6] showed, through retrospective data analysis, that Indomethacin drastically reduces the need for hospitalization. Two recent studies [15,16] have shown the effectiveness of Indomethacin in treating a few SARS-Cov-2 patients with severe co-morbidities. However the sample size in these studies is small and a larger well-planned study was required to validate these findings. This study stems from such a requirement.

## 2.0 Materials & Methods

Two centres were identified for the clinical trial. In both the centres (Narayana Medical College, Nellore, Andhra Pradesh, India, and Datta Meghe Institute of Medical Sciences, Wardha, Maharashtra, India) patients who tested RT-PCR positive for Covid-19 were recruited for an open labelled single arm study for the efficacy and safety of Indomethacin after obtaining Ethics Committee clearance and consent from the patients. Though a double arm Randomized Clinical Trial would have been ideal, we felt that ethically it is unfair to deny patients Indomethacin as our pilot study had shown positive results [15,16]. Patients who opted for Indomethacin were recruited and in order to have a control arm, patients who opted out of Indomethacin and who received paracetamol instead were also monitored with the same blood tests and for other clinical parameters. Though technically it is not a double arm randomized clinical trial, it is now accepted that Propensity Score Matching [17] mimics Randomized Control Trial (RCT). Hence, Propensity Score Matching was applied to match patients in these two arms. The two sets of patients were treated in the same ward of the hospital, by the same set of physicians and during the same period.

Indomethacin replaced paracetamol and was given along with standard care which included hydroxychloroquine, Ivermectin, Azithromycin and vitamins. If patients developed hypoxia, and if the clinician felt the need, they were shifted to a corticosteroid-based regimen. One of the key factors in the study is the development of hypoxia. The standard care drug regimen was a protocol assigned by the Indian Council of Medical Research and it was mandatory for both the arms to follow this regimen. Nevertheless, other studies have shown that hydroxychloroquine and Ivermectin have variable effect in treating Covid-19 patients [18, 19]. A total of 82 patients were recruited from both the centres (75 patients from the first and 7 from the second). In the second centre, 75mg SR (Sustained Release) Indomethacin was administered due to non-availability of 25mg. These cases are called ‘mild and moderate’ in this study, according to WHO clinical progression score (score 4 – hospitalized moderate disease). A total of 109 hospitalized patients on paracetamol instead of Indomethacin formed the control arm. In the second centre, an additional 22 patients, considered as ‘severe’ cases, were recruited. Twenty-one of them were administered supplementary oxygen on admission and one patient required supplementary oxygen subsequently. Though, according to WHO score, an ordinal score 6 (high flow oxygen) is severe, many of the patients were in high flow oxygen on the second day. Hence, all the patients in this group are called severe in this study. These patients were treated with Indomethacin 75mg SR for five days along with Remdesivir (as part of the standard treatment). They were analysed separately as a single arm with the end point being deterioration to a score of 7.

The following were the investigations conducted on admission: CT scan of the lungs, Liver Function Test, Kidney Function Test, C-Reactive Protein and D-Dimer. The blood chemistry was repeated on discharge and the well-being of the patients monitored for fourteen days. The patients were monitored for oxygen saturation, fever, cough and myalgia during the five-day treatment or till recovery. The patients were deemed to have recovered symptomatically if the temperature dropped below 99^0^F for two days and cough reduced to score 2 on a one-to-ten scale (1 – no cough, 2-3 – cough sometimes, 4-6 – cough with the ability to do things, 7-8 persistent cough, and 9-10 great deal of discomfort). Myalgia was left to the patient discretion to report and the patient was discharged with a consistent SpO_2_ value of above 94.

Propensity Score Matching was carried out for the first set of mild and moderate patients using the open-source software R. Age, gender, comorbidities (hypertension, diabetes or both), CT-score (out of 40) on admission, C-Reactive protein on admission, presence or absence of dyspnea were considered as covariates. A logistic regression function with Iteratively Weighted Least Square, the default algorithm in R for the command ‘glm’, was used to identify the weights. The Hosmer and Lemeshow goodness of fit test returned a p-value of 0.729, thus ascertaining a good fit. Fischer scoring algorithm converged in 4 iterations and the deviance check also confirmed a good fit. Propensity Scores were calculated and the matching carried out using ‘MatchIt’ command of R. A greedy algorithm with ‘nearest’ as the option for search and a caliper of 0.12 was arrived at by trial and error. Out of a total of 82 patients in the Indomethacin arm, 72 patients were matched with the patients from the paracetamol group, which had 109 patients. In order to understand the impact of the sample size, the response rate for paracetamol was assumed to be 0.8 and that for Indomethacin to be 0.96. The sample size was calculated using R with an alpha value of 0.05. Marginal power was 0.84, above the minimum recommended limit of 0.8 [20]. Post-hoc calculations based on the actual result gave a marginal power of 0.99.

The calculated propensity scores for Indomethacin and paracetamol groups are shown in Fig. 1. A good match of propensity scores is evident.

**Fig. 1.**
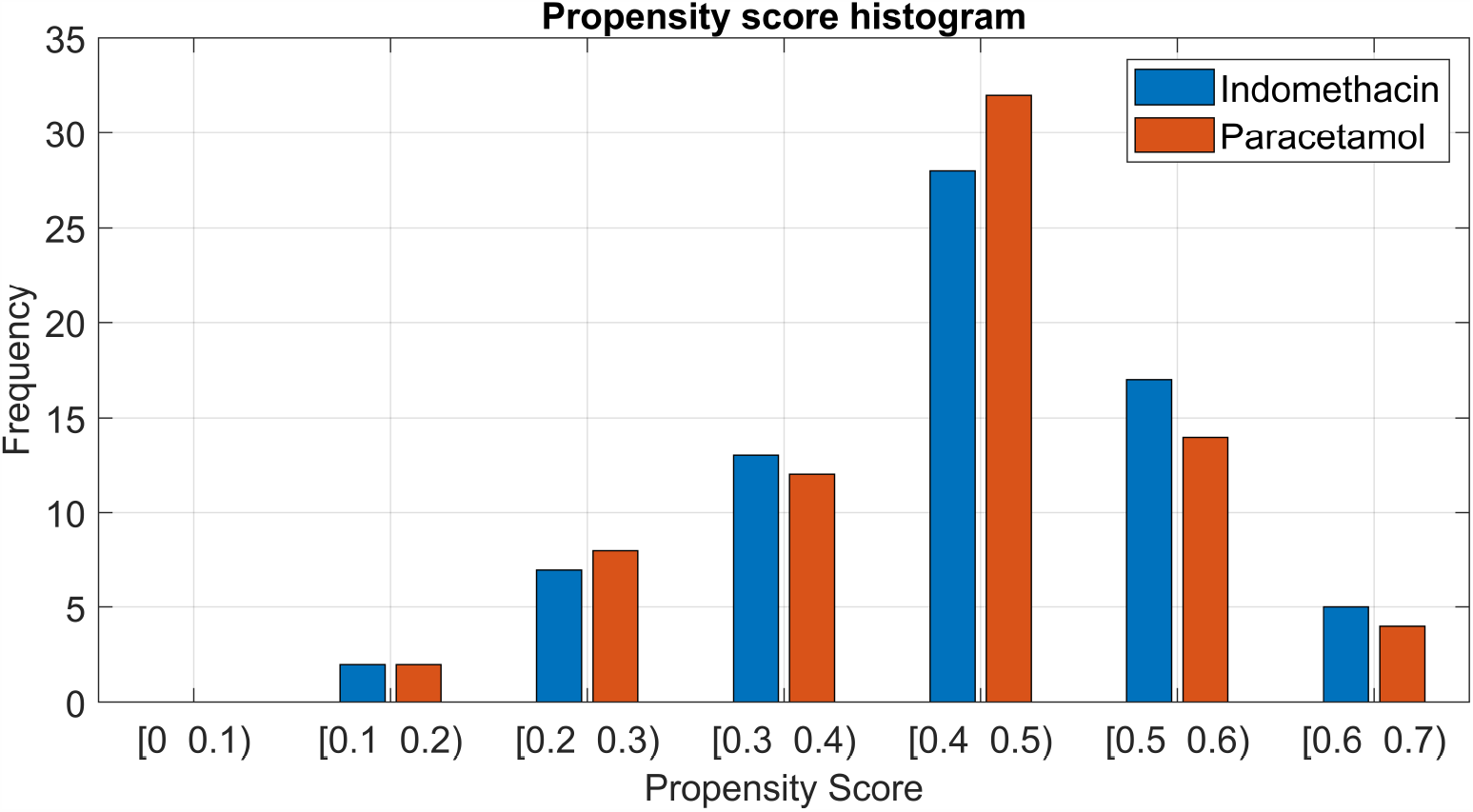
Matched Propensity score for Indomethacin and Paracetamol arm

Balance plots in Fig. 2 for all the covariates and the jitter plot in Fig. 3 bring out clearly the relation between the control and treatment group after matching.

**Fig. 2a.**
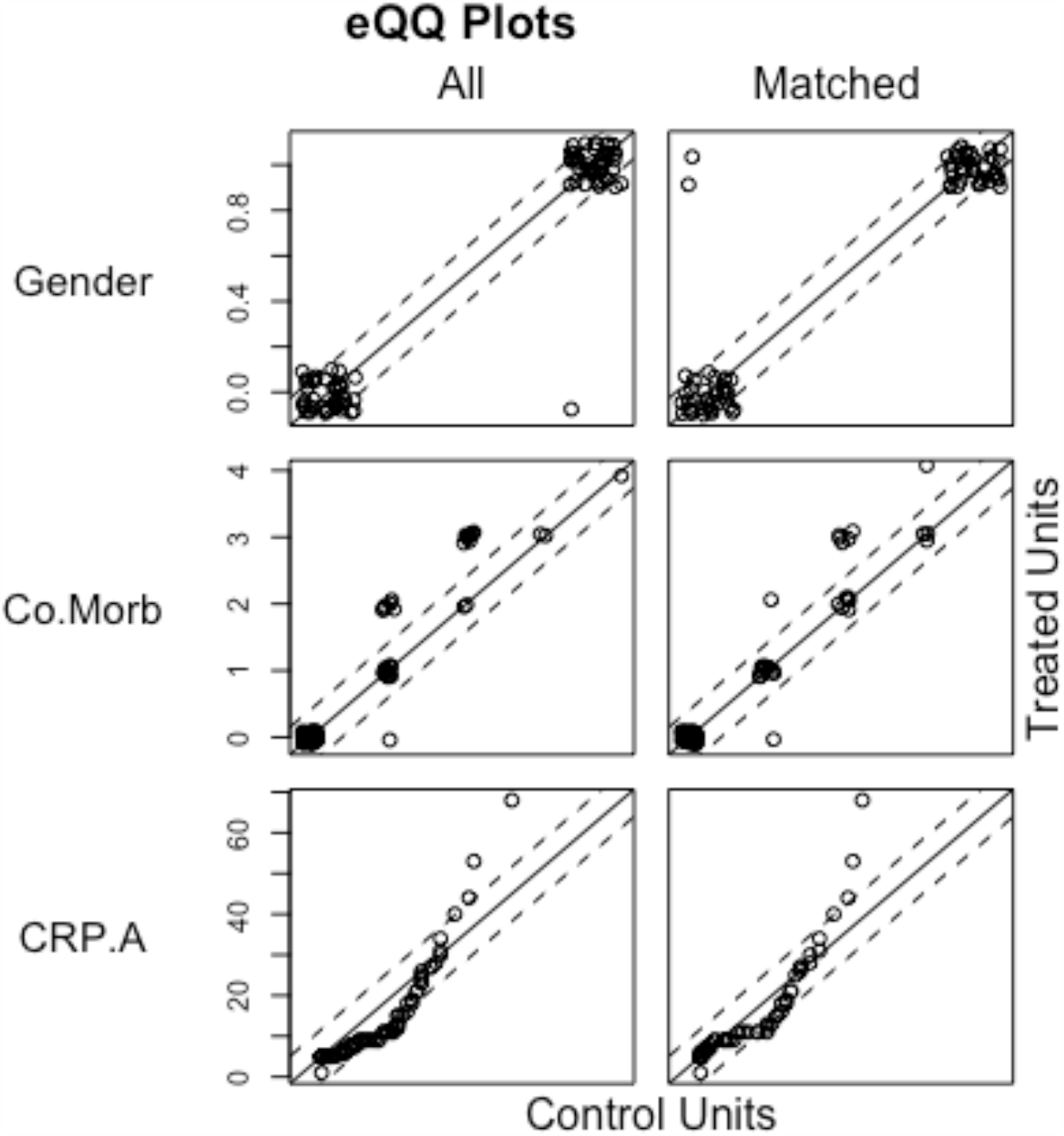
Balance Chart for Covariates Gender, Co-Morbidity and CRP-A

**Fig. 2b.**
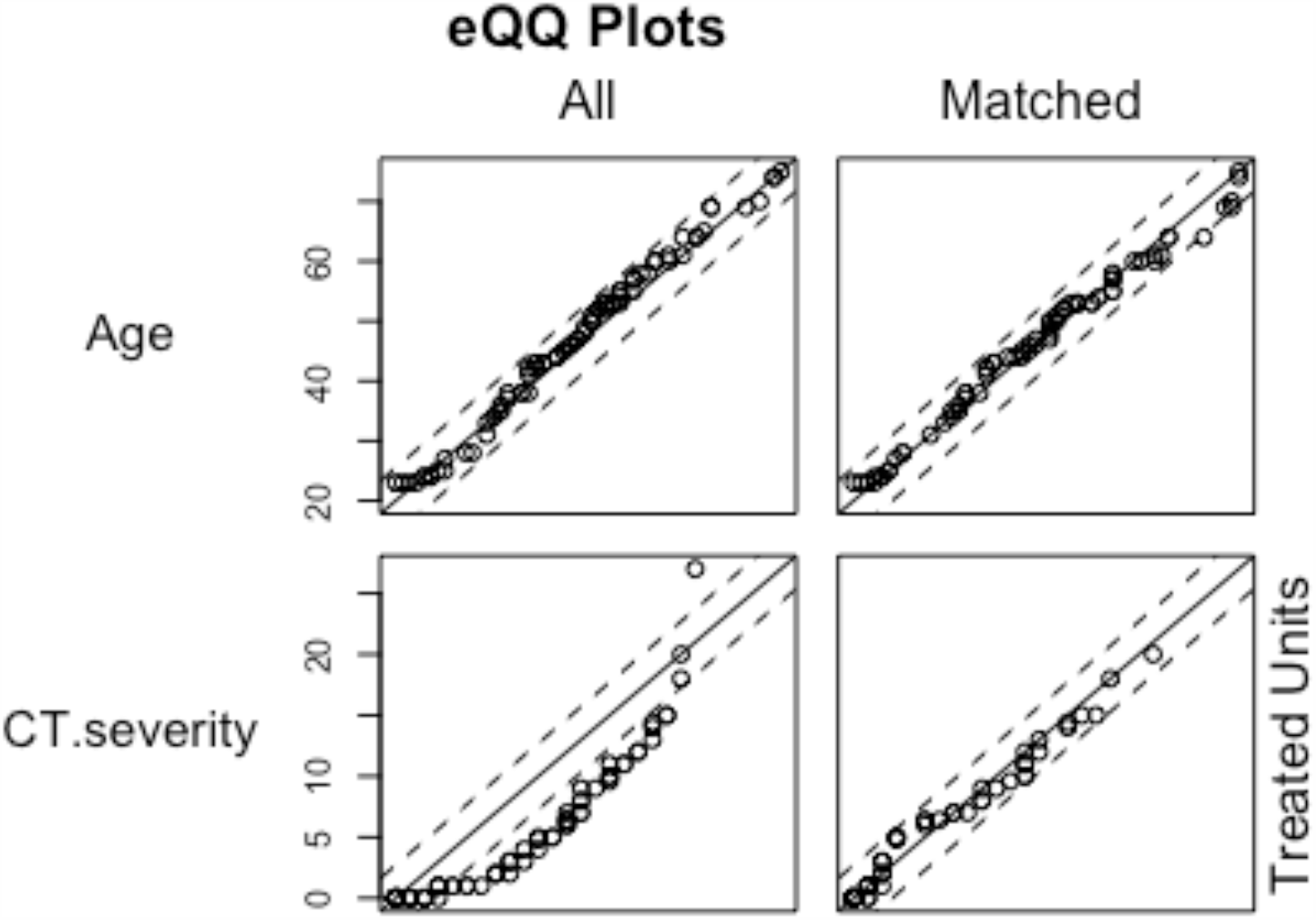
Balance Chart for Covariates Age and CT Score

**Fig. 3.**
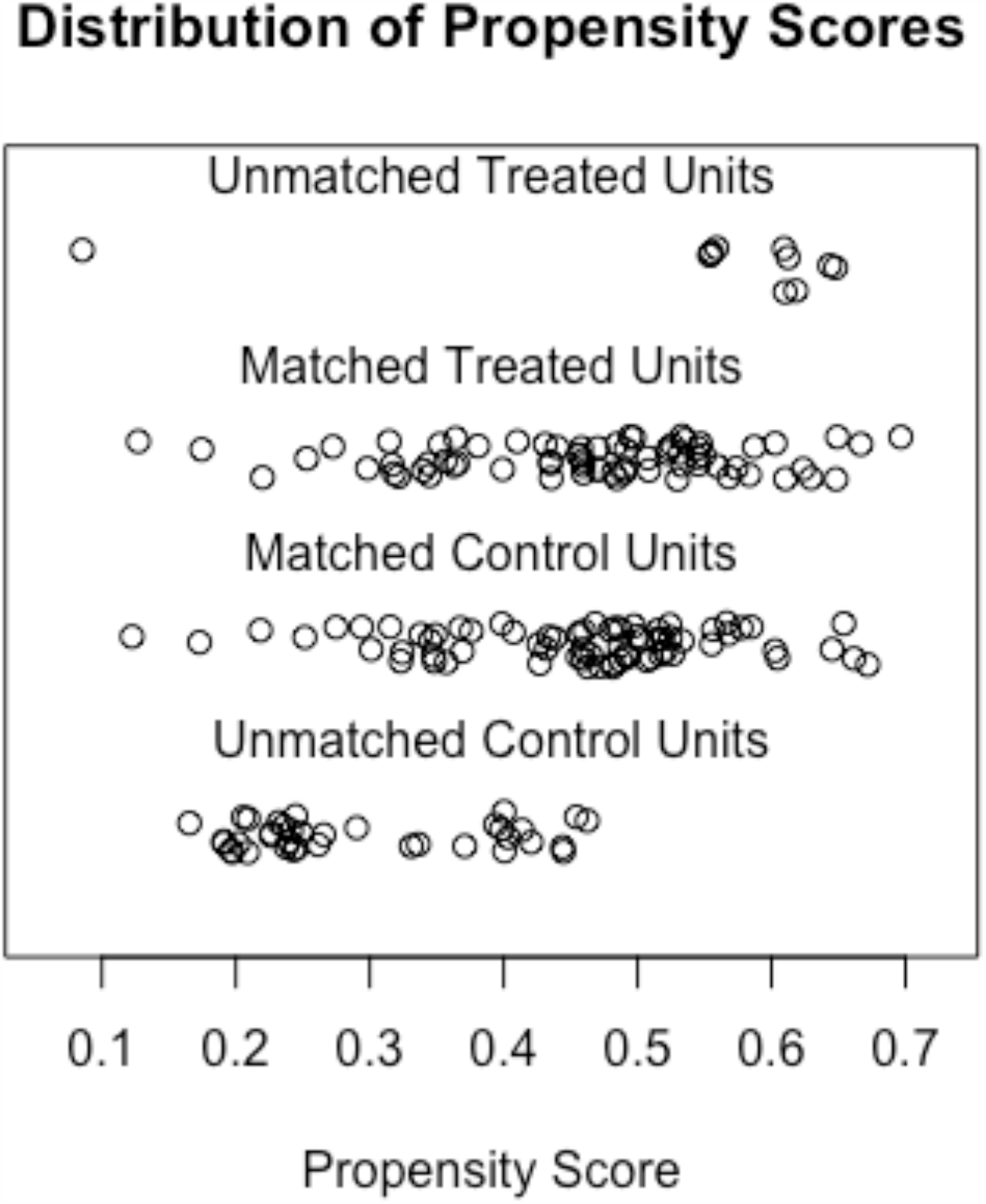
Jitter plot of the propensity score.

In order to further understand the details of the matched patient profiles and that of the severe patients, various covariates are replotted in Figs 4 to 7. Fig. 4 is the age and co-morbidity profile, Fig 5 and Fig.6 give details of the CT score and Fig 7 shows the CRP distribution of the patients. A close match of the profiles between the two arms is evident from these figures. The CRP and the CT scores of severe patients reveal the extent of the disease.

**Fig. 4.**
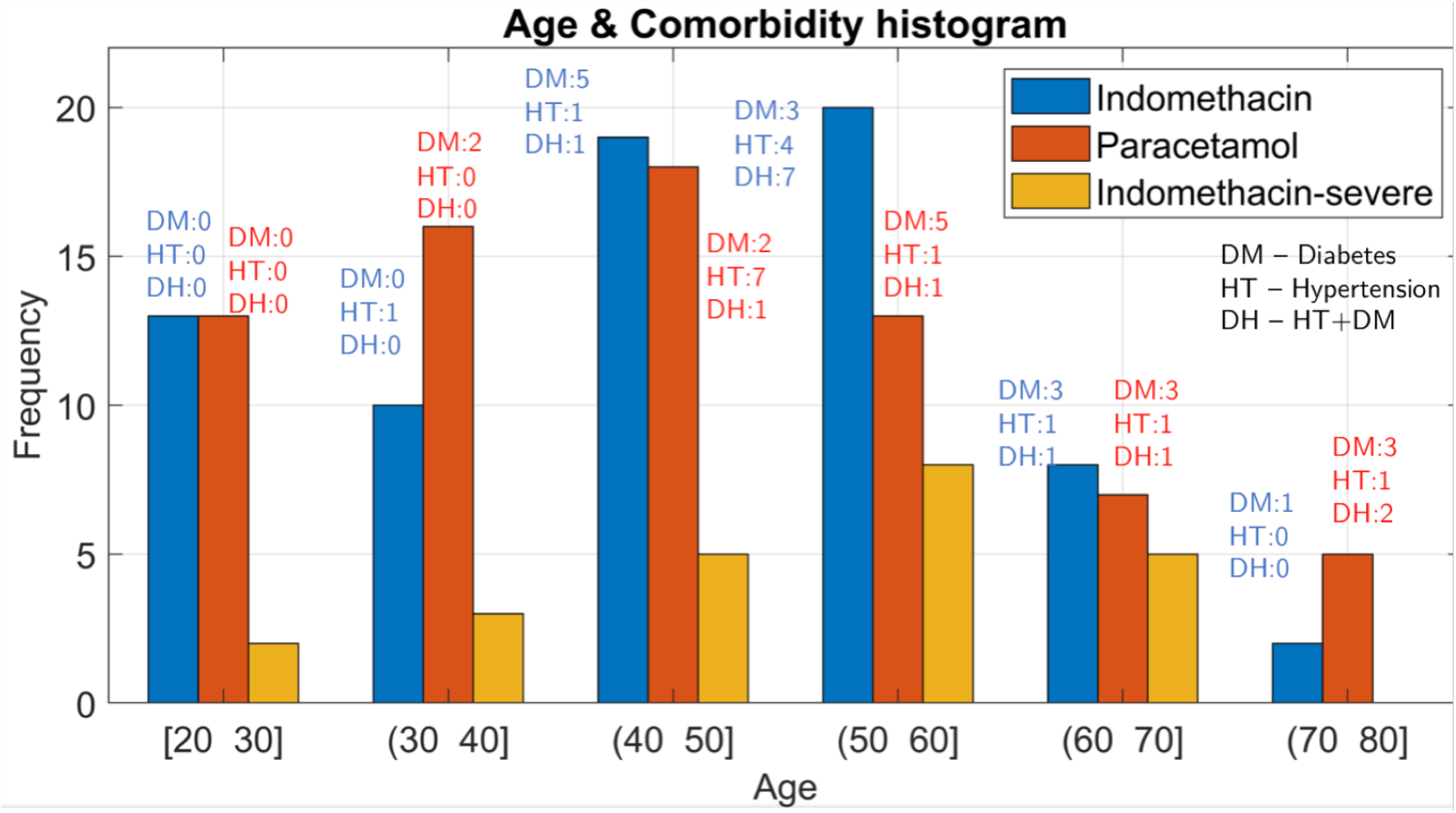
Age and Comorbidity profile of various arms

**Fig. 5.**
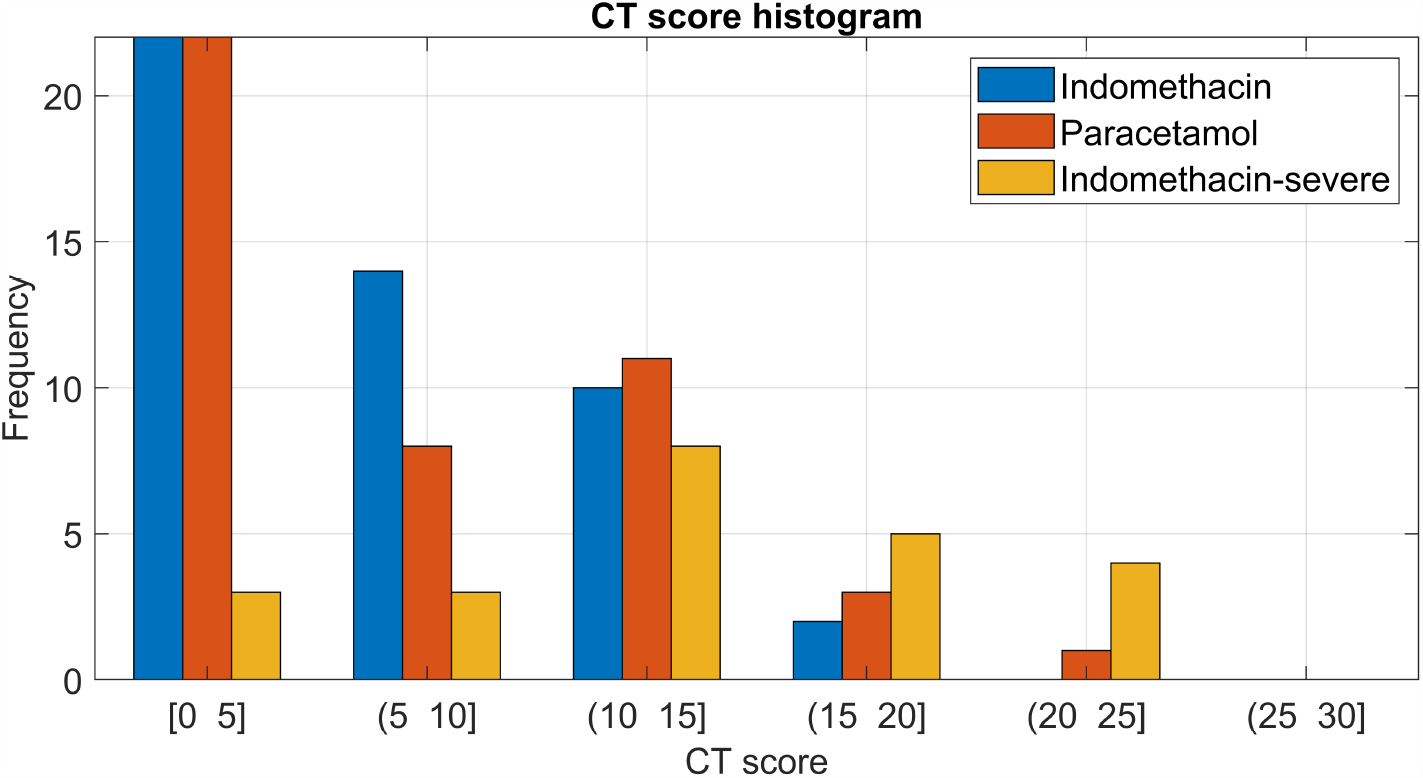
CT – Score profile of the patients

**Fig. 6.**
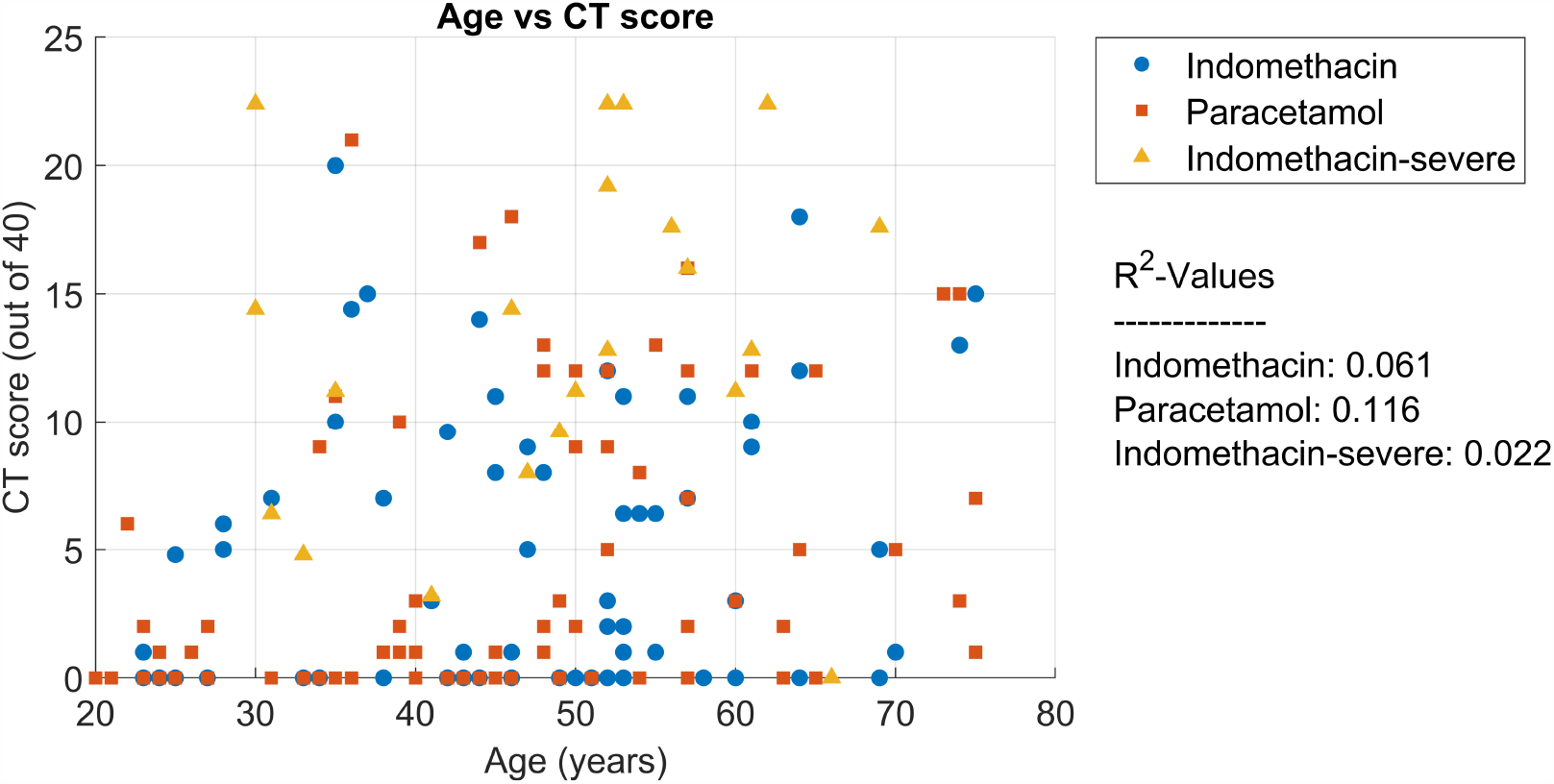
CT-Score (out of 40) on admission as a function of Age

**Fig 7.**
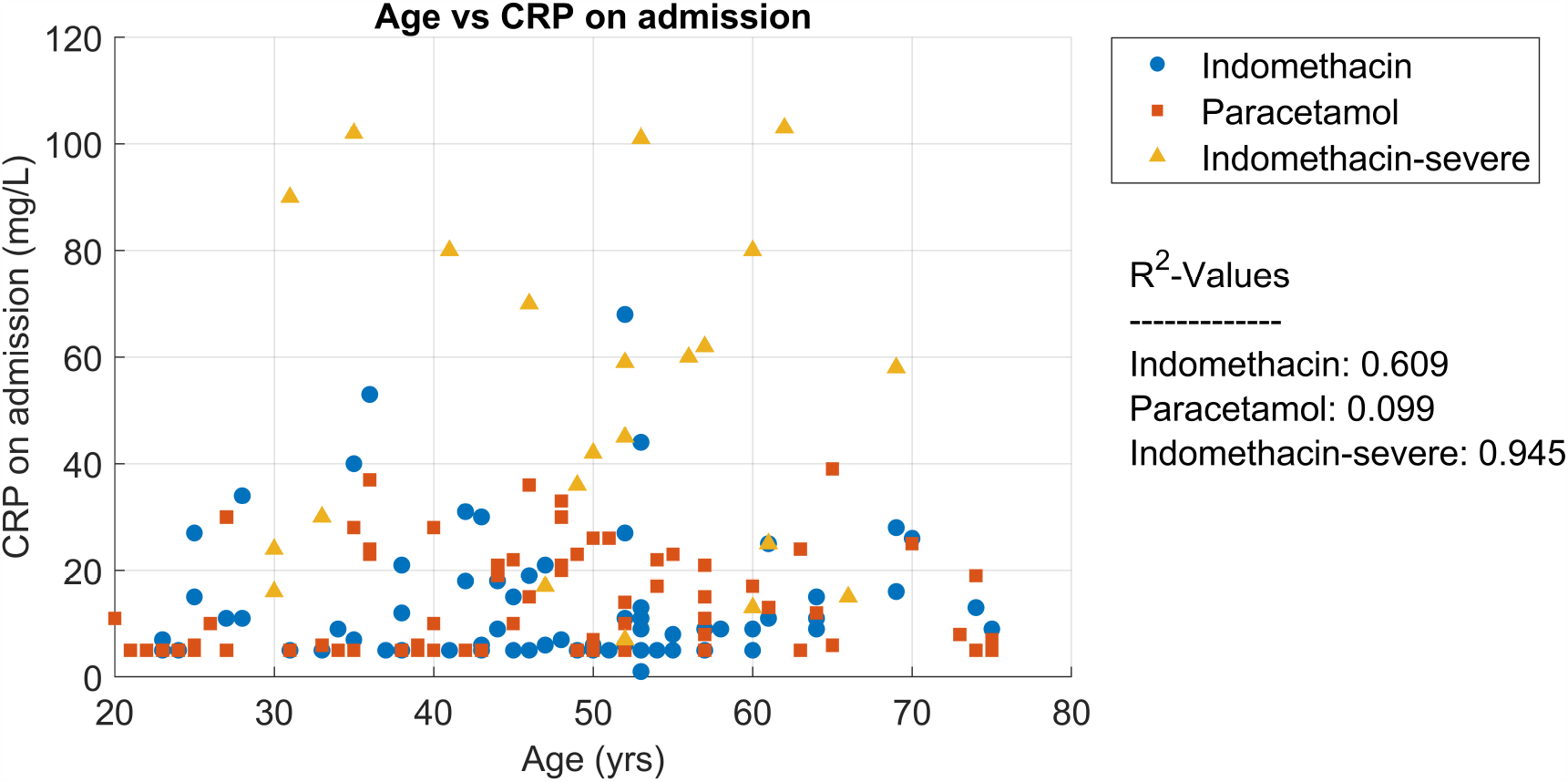
CRP distribution on admission as a function of age

## 3.0 Results

### 3.1 Efficacy of Indomethacin

Indomethacin was hypothesized to be associated with symptomatic relief, namely the number of days for becoming afebrile, days for reduction of cough to two in an ordinal scale (occasional) and relief from myalgia. These were monitored and the results are shown in Fig. 8. The symptomatic recovery from fever, cough and cold in terms of median values is shown in Table 1. The results are from a one-sample Wilcoxon test and IQR indicates InterQuartile. Range. The Table clearly brings out the recovery in the Indomethacin arm of the study.

**Table 1.** Symptomatic Relief due to various treatments

| Treatment | Days to become Afebrile |  |  | Days for Cough Reduction |  |  | Days for Myalgia Reduction |  |  |
| --- | --- | --- | --- | --- | --- | --- | --- | --- | --- |
|  | Median | 95%CI | IQR | Median | 95%CI | IQR | Median | 95%CI | IQR |
| Indomethacin- Mild and Moderate | 4 | 3.5,4.0 | 1 | 3 | 3.0,3.5 | 2 | 3 | 3.0,4.0 | 2 |
| Paracetamol – Mild and Moderate | 7 | 7.0,8.0 | 1 | 8 | 7.0,7.5 | 2 | 6.5 | 5.5,7.5 | 3.25 |
| Indomethacin – Severe | 2 | 2.0,2.0 | 0 | 3 | 2.5,3.0 | 0.5 |  |  |  |

**Fig. 8.**
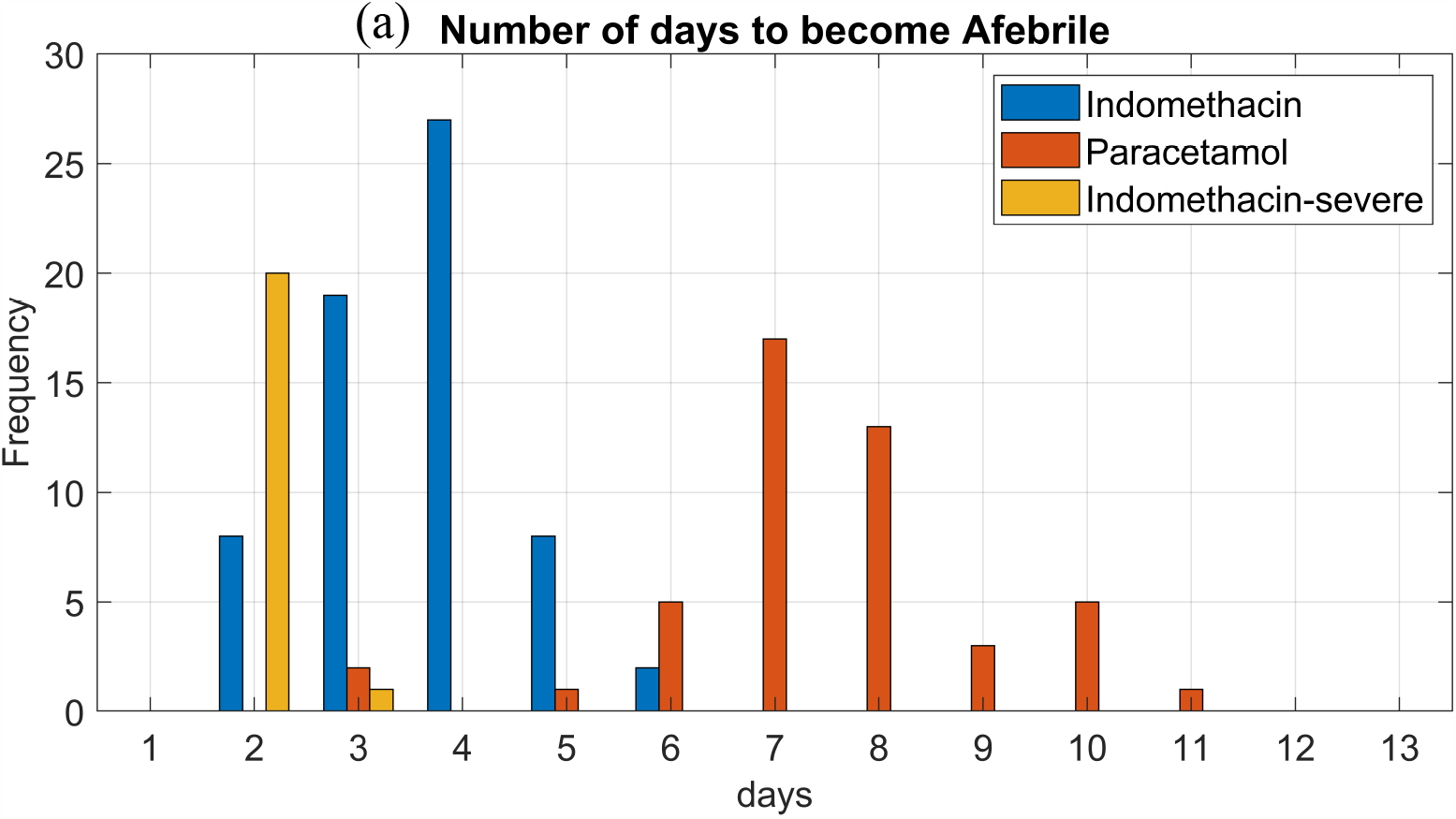
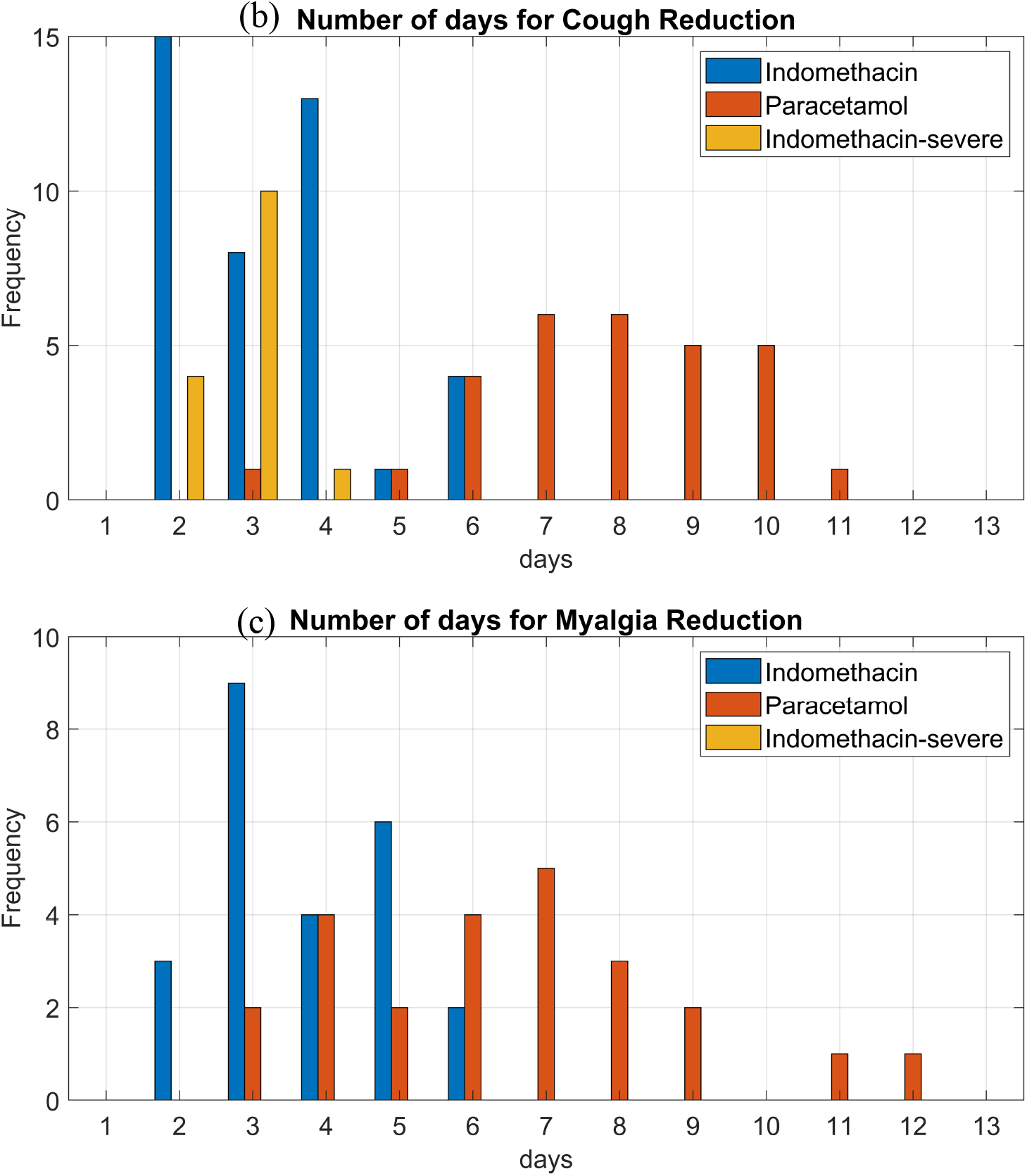
Symptomatic relief of patients with Indomethacin and Paracetamol

In order to rule out the association of temperature on admission, days for becoming afebrile was plotted against temperature on admission and shown in Fig. 9. One can conclude from Figs. 9 and 10 that the temperature on admission or the CT score on admission probably has no relation to the patient recovery.

**Fig. 9.**
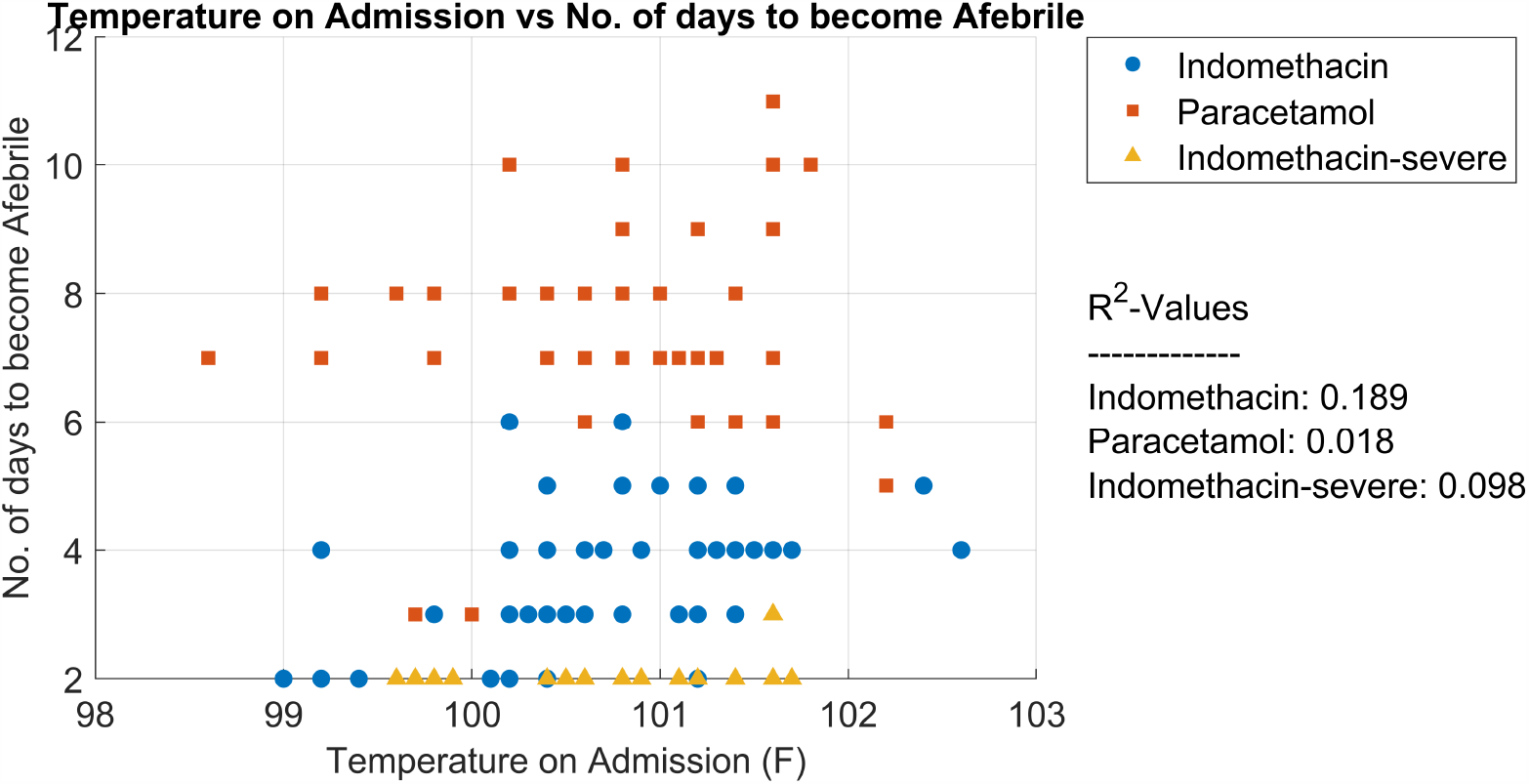
Effect of temperature on admission on days for becoming Afebrile.

**Fig. 10a.**
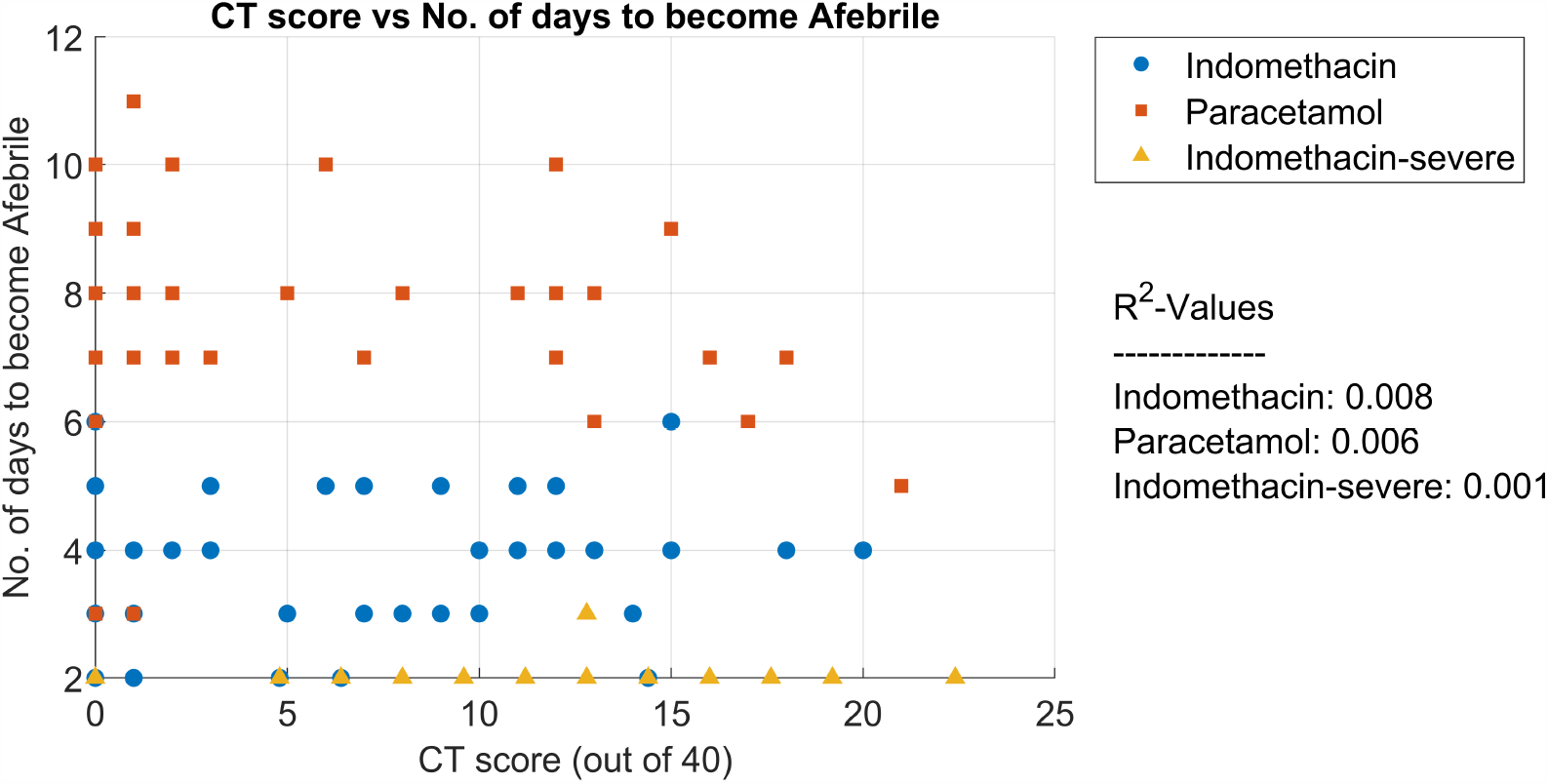
Effect of CT-Score on time for symptomatic relief – Days to become Afebrile

**Fig. 10b.**
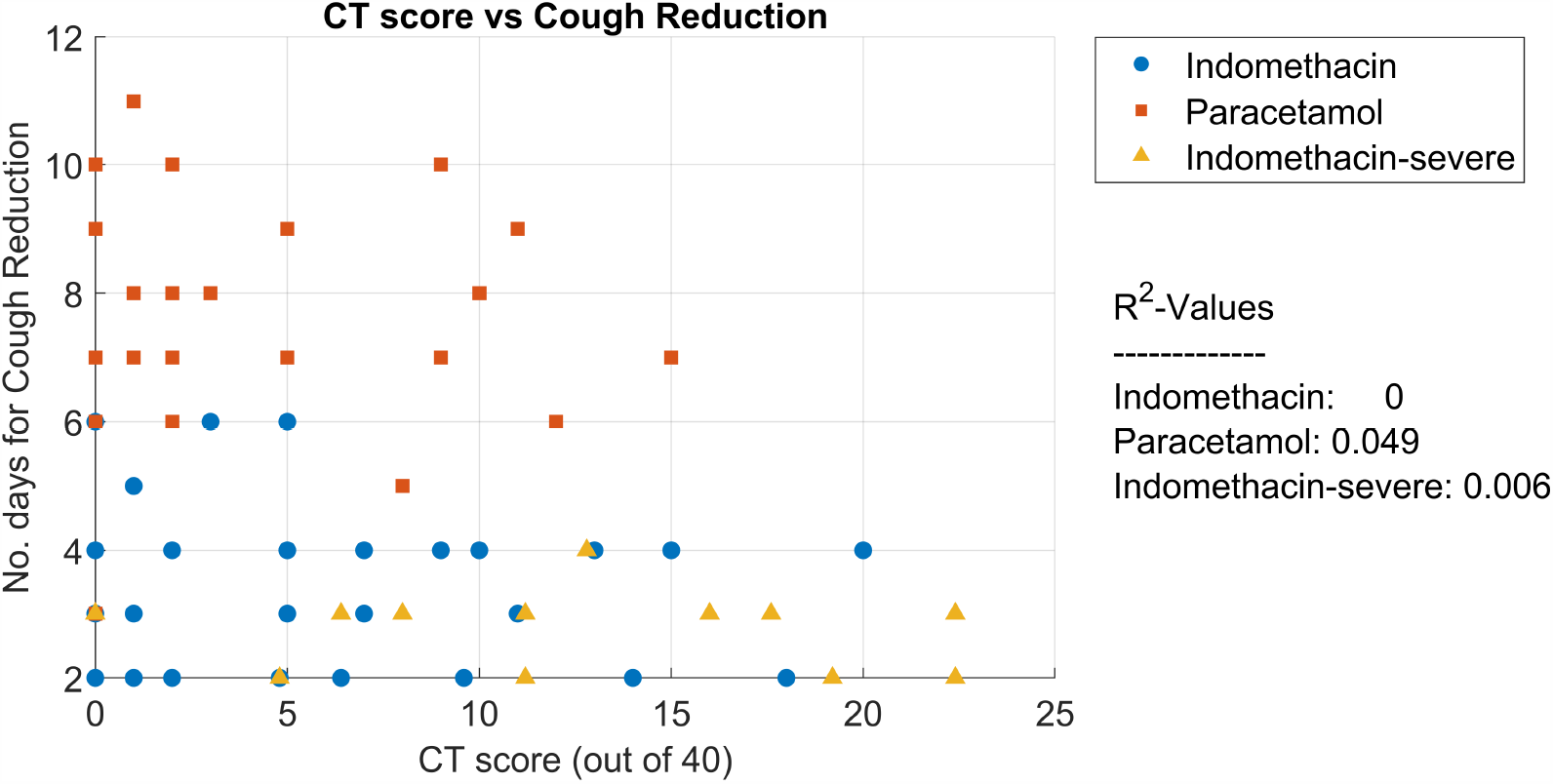
Effect of CT-Score on time for symptomatic relief – Days for Cough reduction

The two key questions in this study are: How many patients developed hypoxia and required steroids treatment, and how many had a prolonged stay (more than 14 days) in the hospital. We split the patients requiring supplementary oxygen into two categories. The first category consisted of patients admitted with shortness of breath and oxygen saturation more than 95% and requiring supplementary oxygen subsequently during the hospital stay due to drop in saturation. This statistic is given in Fig. 11. Interestingly, only one patient in the Indomethacin arm required supplementary oxygen at 2L/min for two days.

**Fig. 11.**
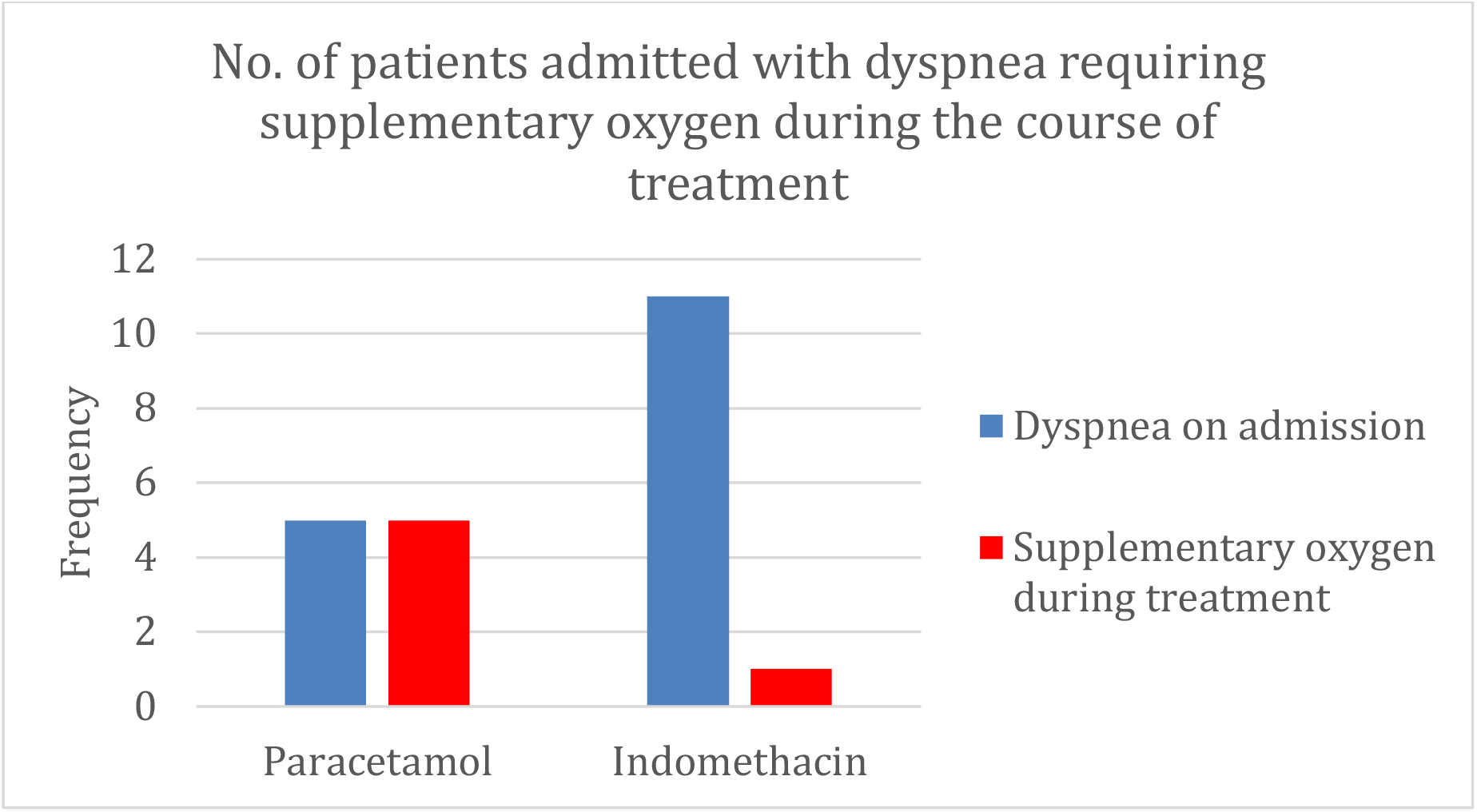
Number of patients admitted with dyspnea and subsequently required supplementary oxygen

The other set is of patients who had no shortness of breath on admission but developed hypoxia during the course of treatment. This is shown in Fig. 12. One patient in the Indomethacin group, who had vomiting and nausea on admission and during treatment, had a brief period of hypoxia but did not require supplementary oxygen.

**Fig. 12.**
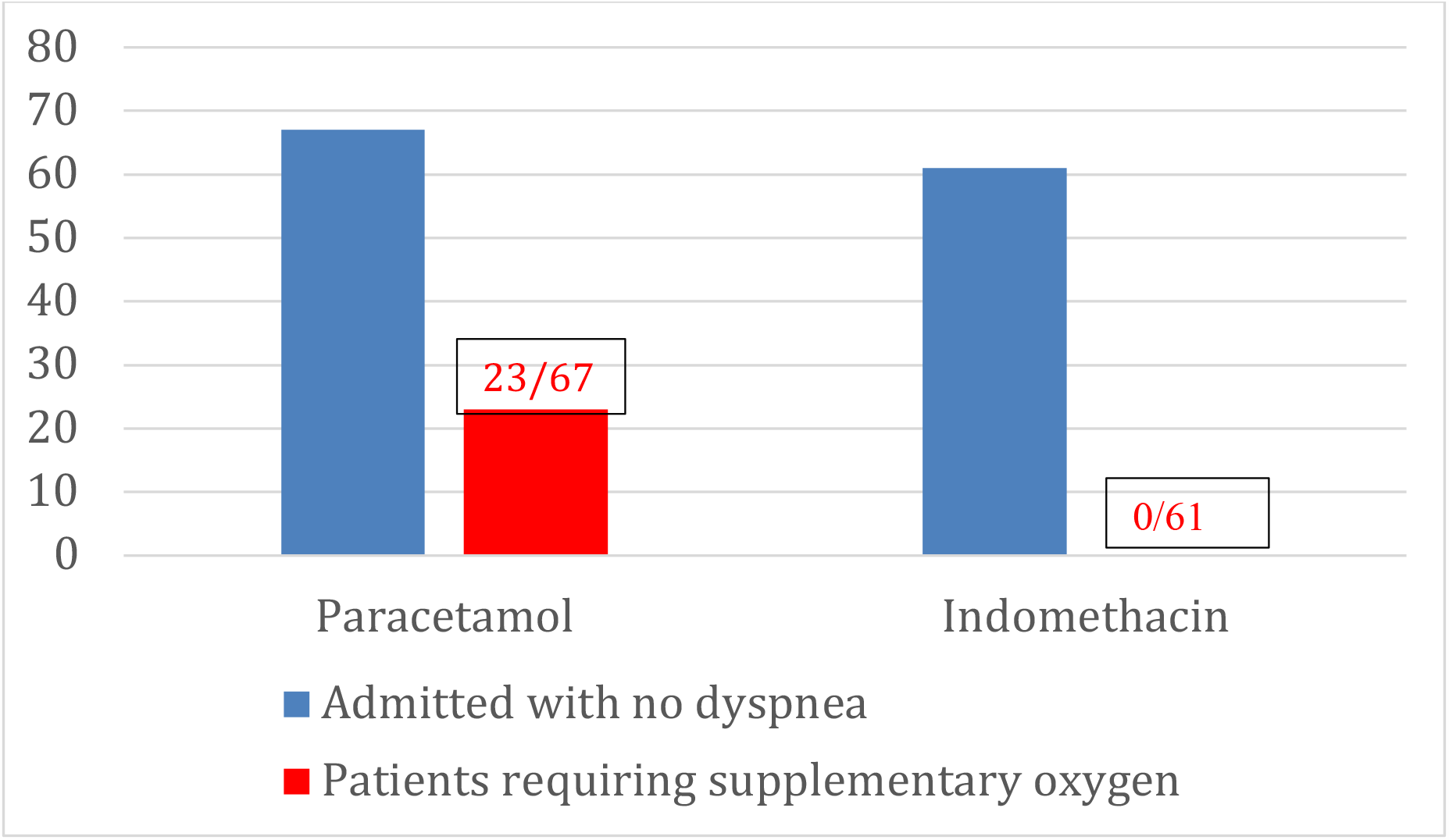
Patients admitted with no dyspnea but subsequently required supplementary oxygen

Fig. 13 gives the probability of developing hypoxia using the Kaplan-Meier survival model. Patients in the paracetamol arm required supplementary oxygen even after four-five days of treatment. The hazard ratio of the Indomethacin arm compared to the paracetamol arm, using the Cox-proportional-hazards model, is a low 0.00218 with a 95 % CI being (0.00015 to 0.03)

**Fig. 13.**
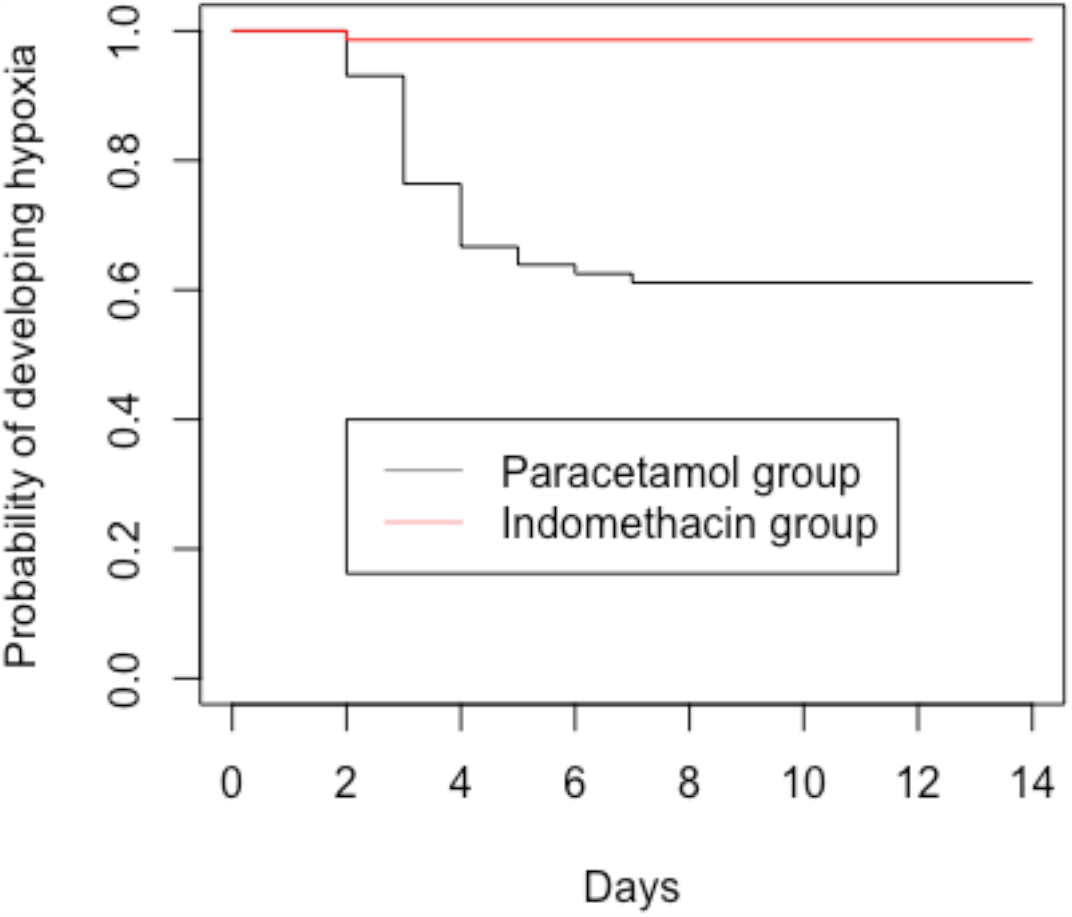
Kaplan-Meier curves for the probability of developing hypoxia

The results from the software R for a Cox proportional hazards model is shown in fig.14. It can be seen that apart from the indomethacin (treatment) and shortness of breath at admission (SOB), CT-score and to a lesser extent C-Reactive protein (CRP.A) have significance for the development of hypoxia. The effect of CT score is further explained by Fig. 15

**Fig. 14.**
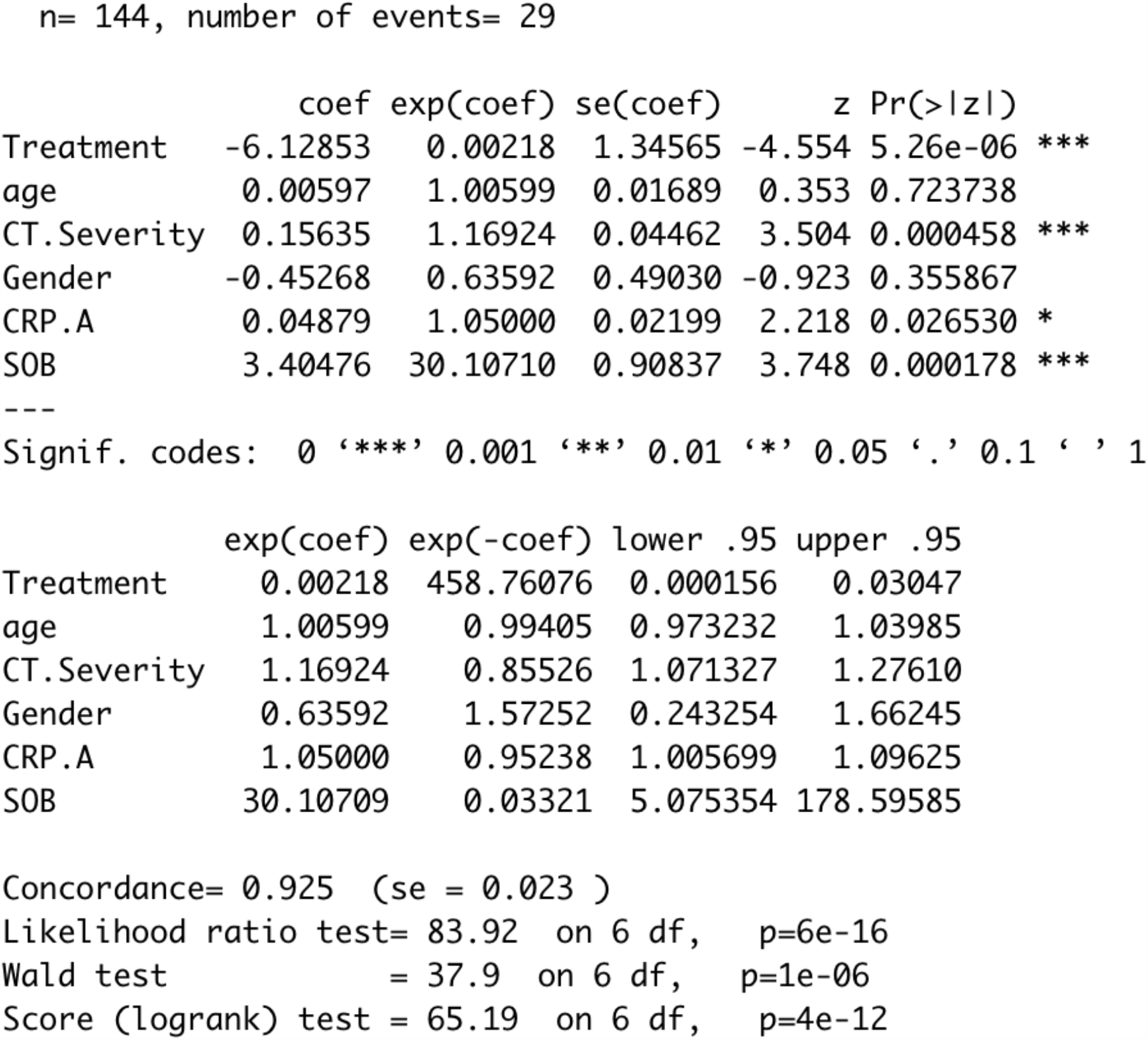
Results of Cox proportional hazards model

**Fig. 15.**
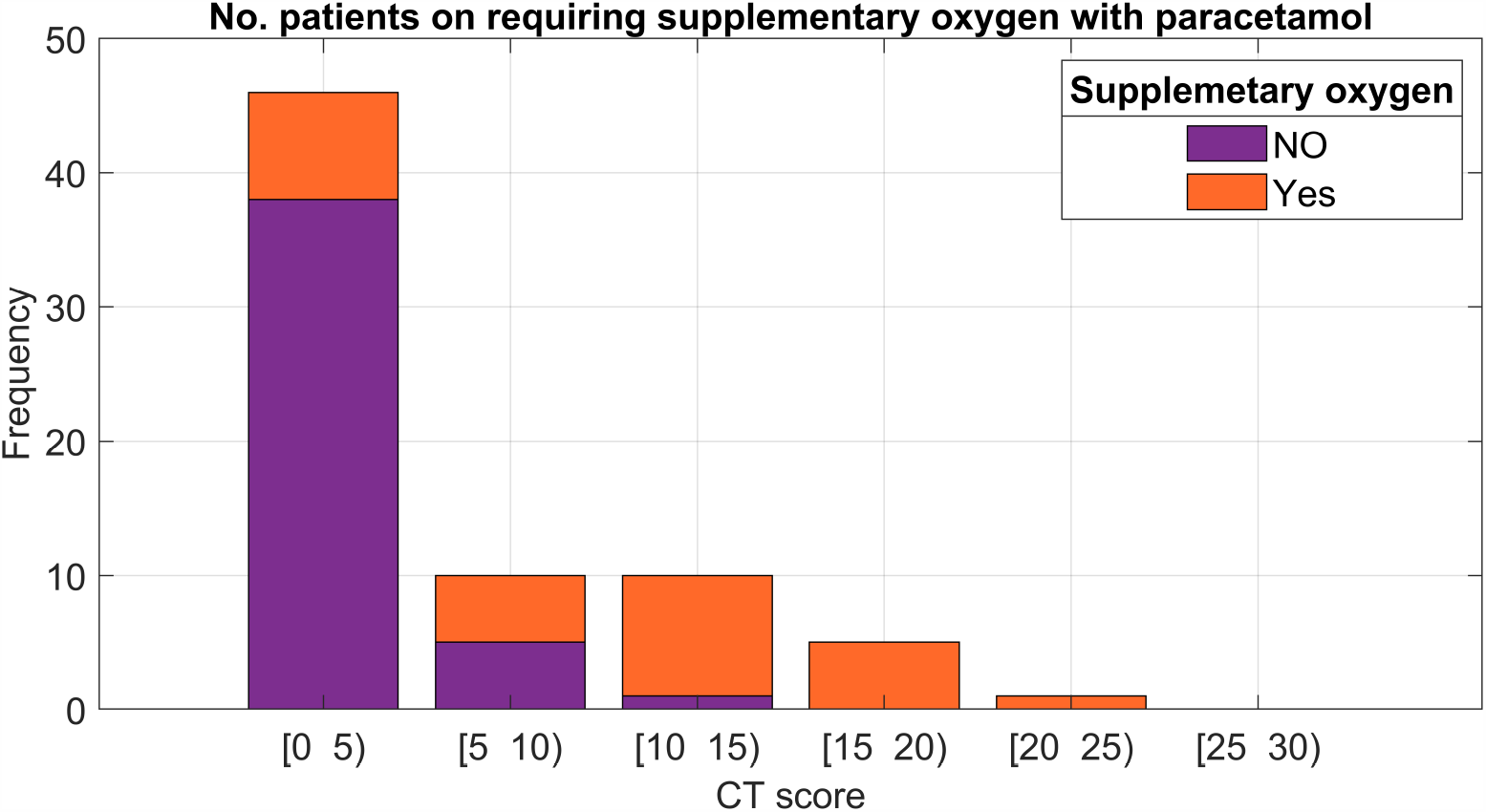
Patients requiring supplementary oxygen as a function of CT score.

Patients with prolonged stay in the hospital in the paracetamol arm were analysed for hazard ratio. Development of hypoxia seems to be the only significant factor with a hazard ratio of 5.2 (95 % CI [1.27 to 21.6].

One of the key roles for Indomethacin is to act as an anti-inflammatory drug. The change in CRP for the Indomethacin arm is given in Fig. 16. The paracetamol arm, in which 39 out of 72 patients were treated with methylprednisolone along with paracetamol, is not included for comparison.

**Fig. 16.**
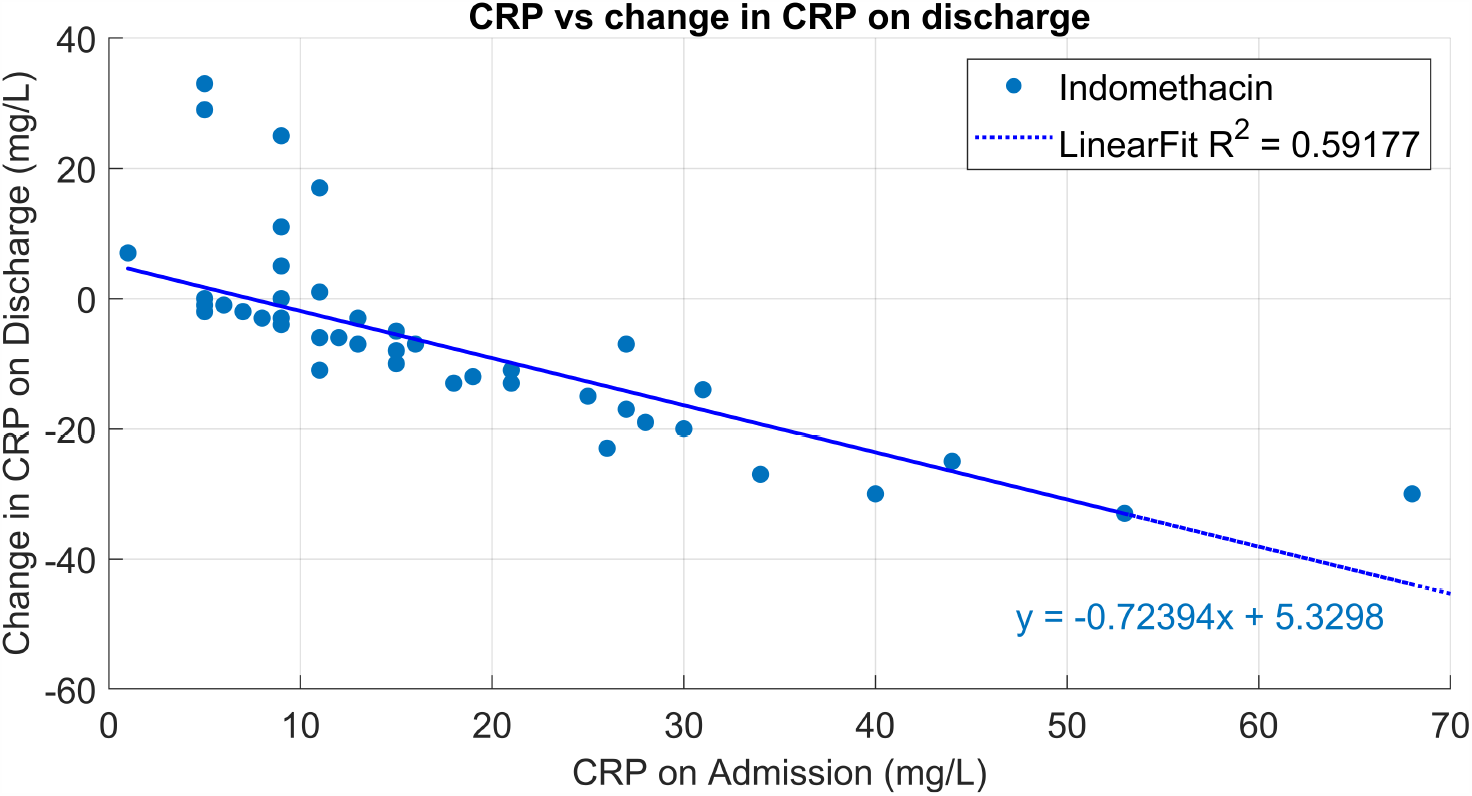
Change in CRP after indomethacin treatment

Patients were followed up after 14 days by telephone since they were discharged early after 6 days in the Indomethacin arm. None of them reported any significant symptom. On the other-hand, most patients of the paracetamol arm were in hospital for ten to fourteen days

A group of 22 patients, as mentioned earlier, on 75mg SR of Indomethacin, with more severe Covid-19, was also monitored in the study. The patients’ profiles are given in the preceding figures. The number of days for recovery to WHO ordinal scale for clinical improvement 4 (no oxygen) are given in Fig. 17. Twenty-one patients were discharged on or before 14 days and one patient, who had acute pancreatitis, was discharged after 17 days. Most importantly, no patient required ICU admission. We did not compare this group to a similar one with paracetamol.

**Fig. 17.**
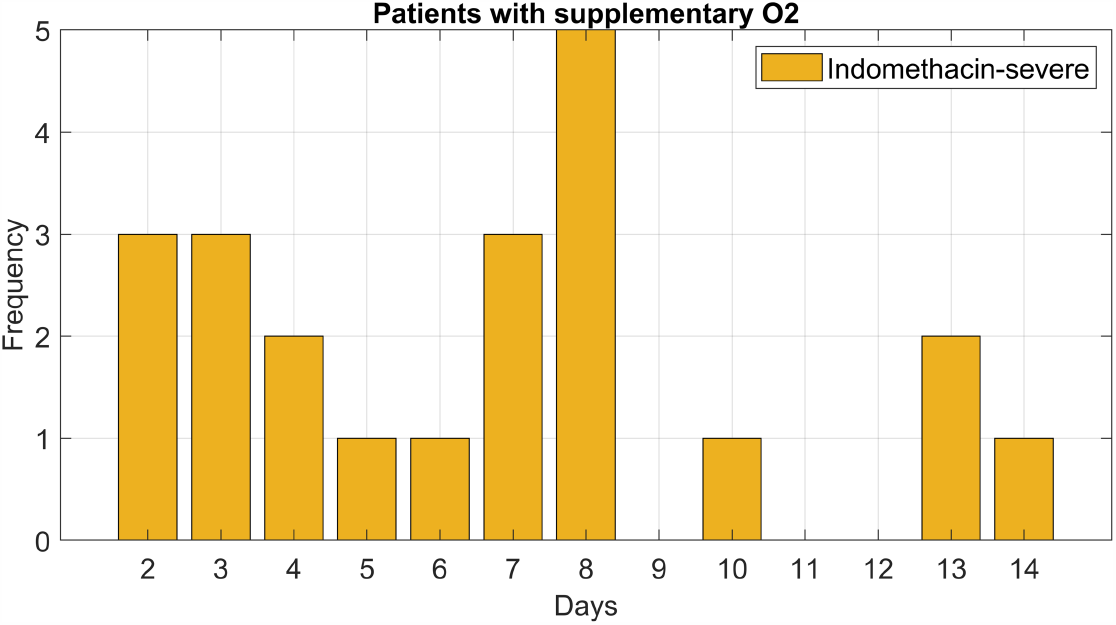
Number of days to wean away the patients from supplementary oxygen

The reduction of C-Reactive Protein for these patients is plotted in Fig. 18. The reduction in CRP with Indomethacin is similar to that of mild/moderate patients.

**Fig. 18.**
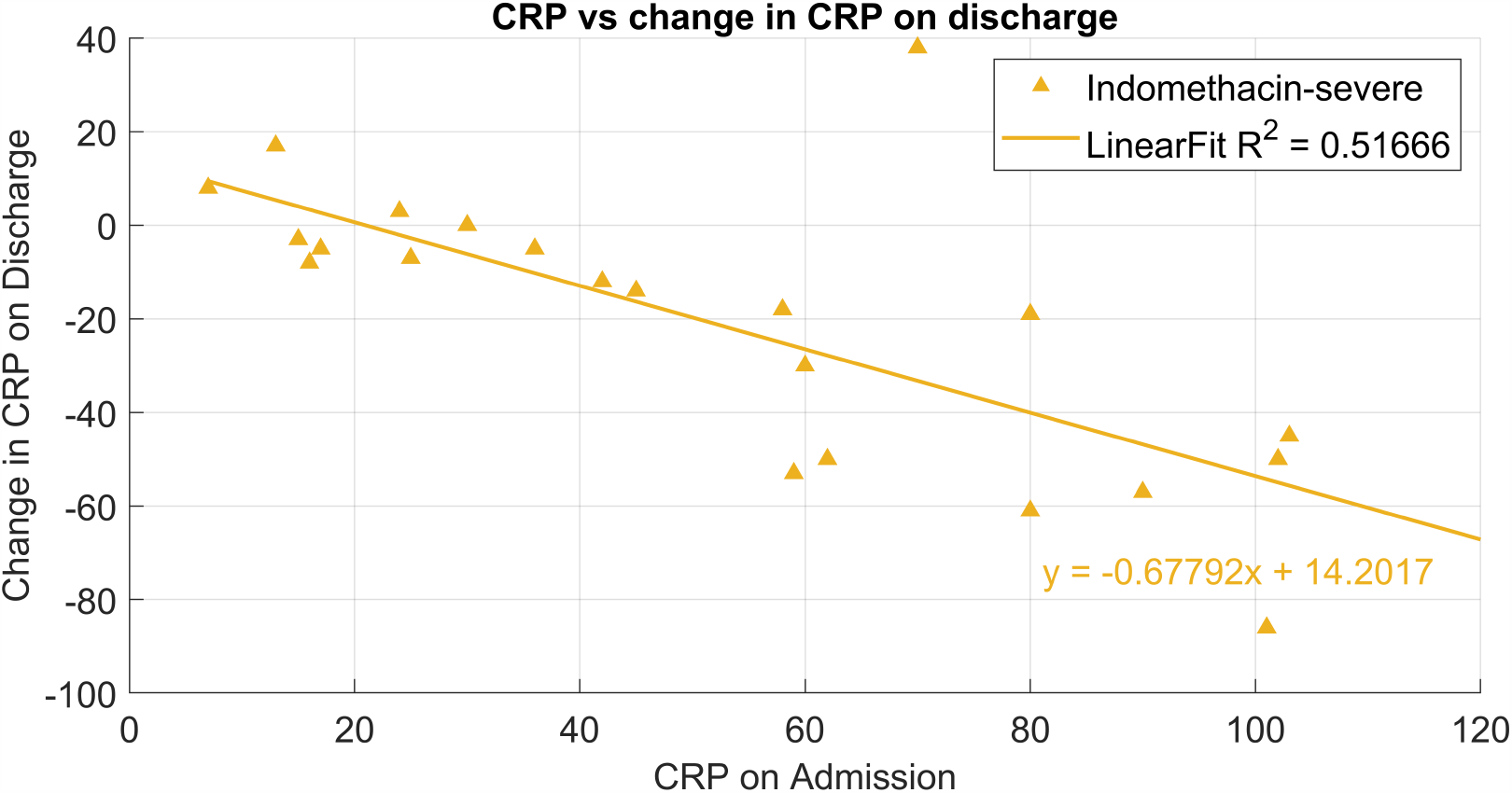
Reduction of CRP in severe patients

### 3.2 Safety profile of Indomethacin

Though Indomethacin was approved in 1965, there have been concerns about its safety [21]. Patients were tested for Serum Urea and Creatinine, SGOT and SGPT before and after the treatment and the results are given in Figs 19-22.

**Fig. 19.**
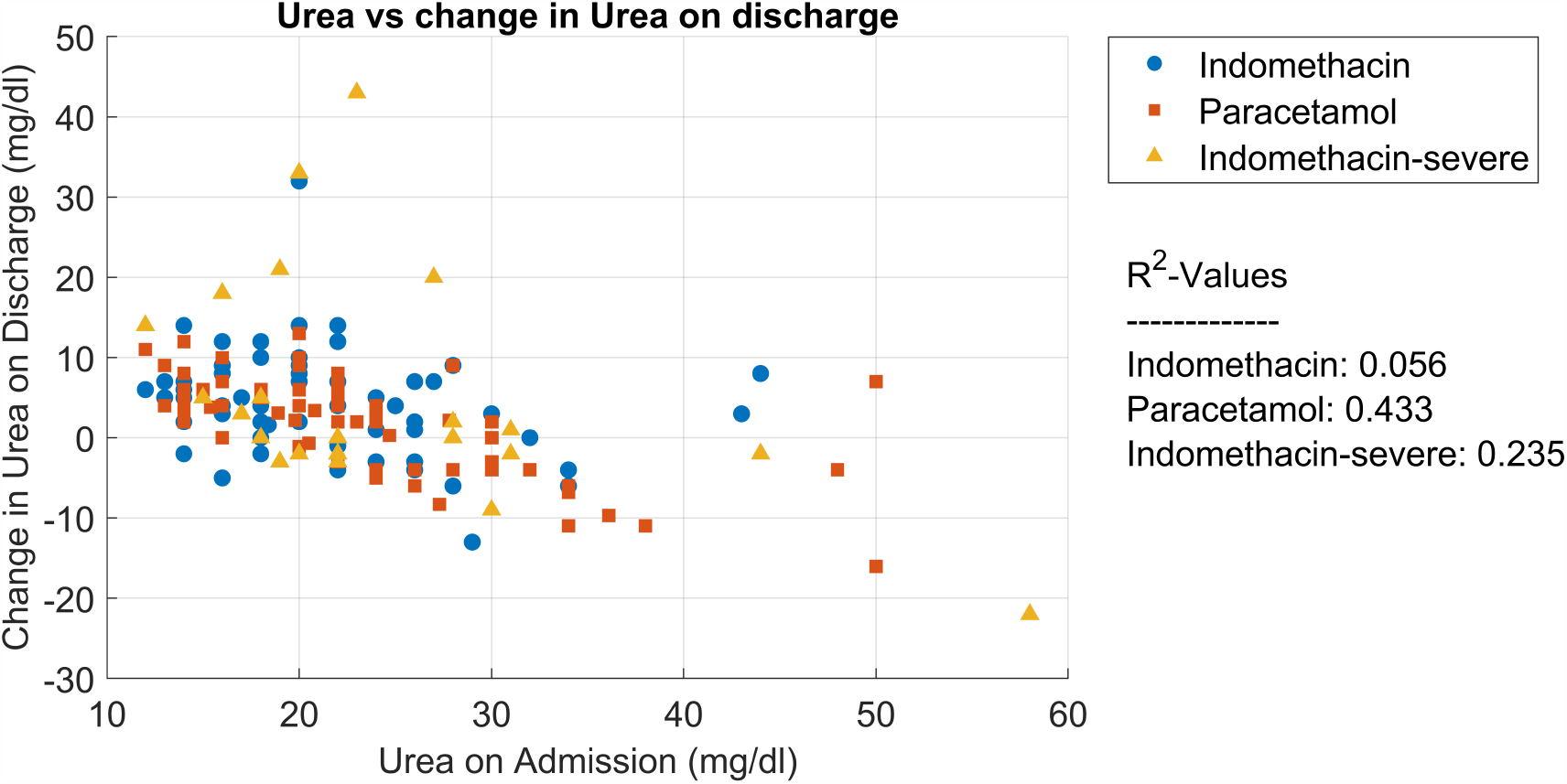
Change in Serum urea after treatment as a function of urea on admission

**Fig. 20.**
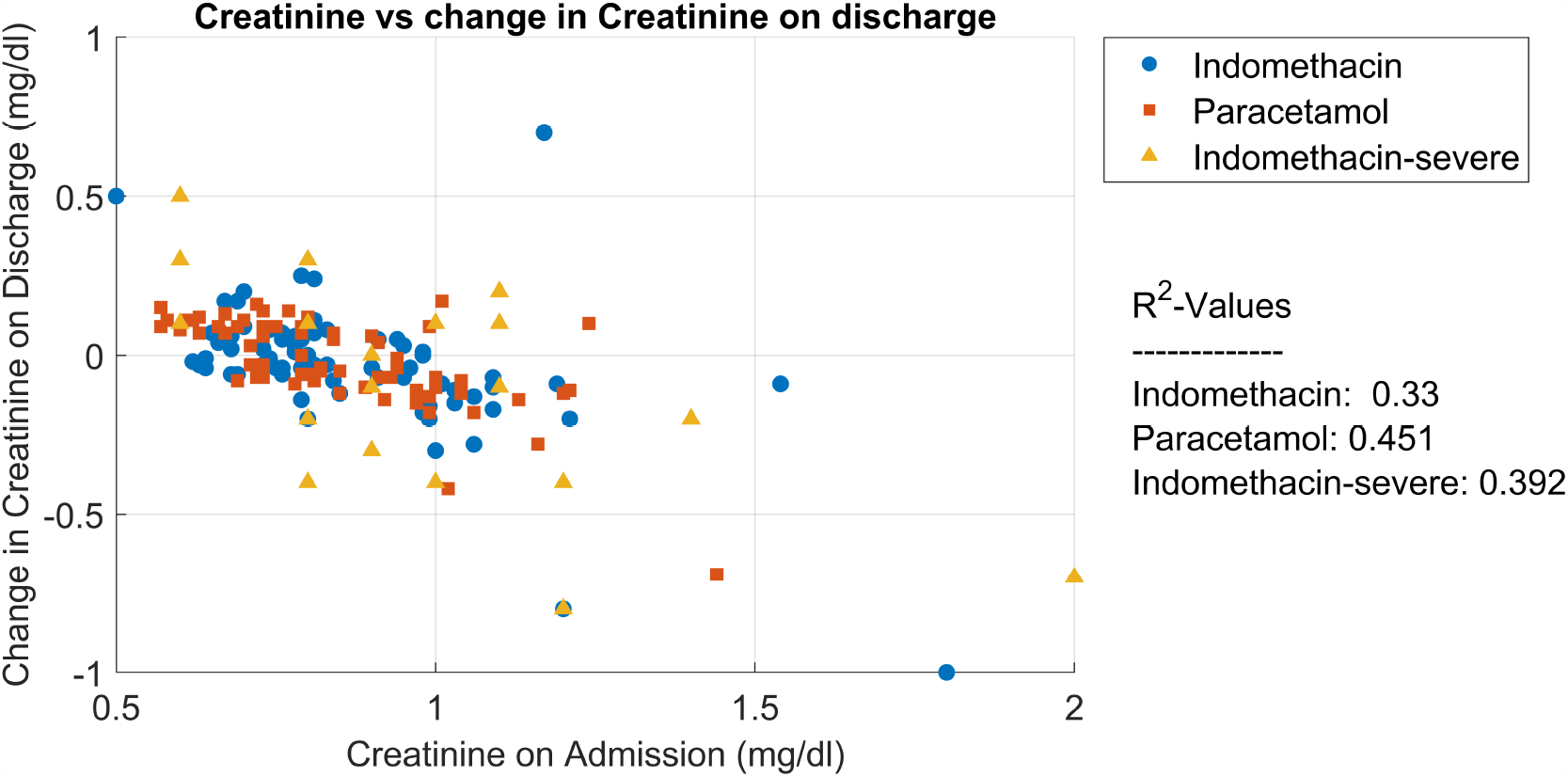
Change in Serum Creatinine after treatment as a function of creatinine on admission

**Fig. 21.**
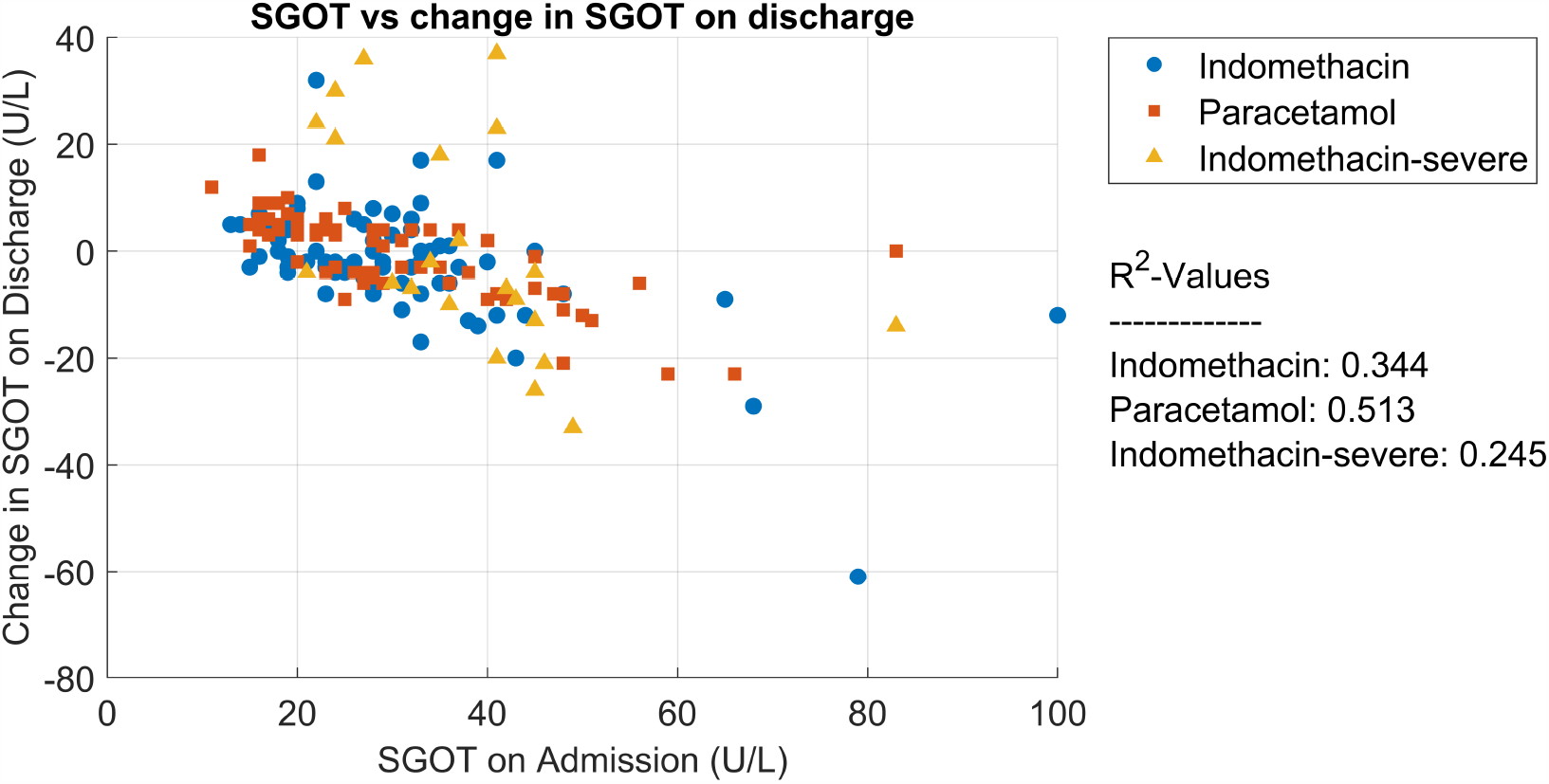
Change in SGOT after treatment as a function of SGOT on admission

**Fig. 22.**
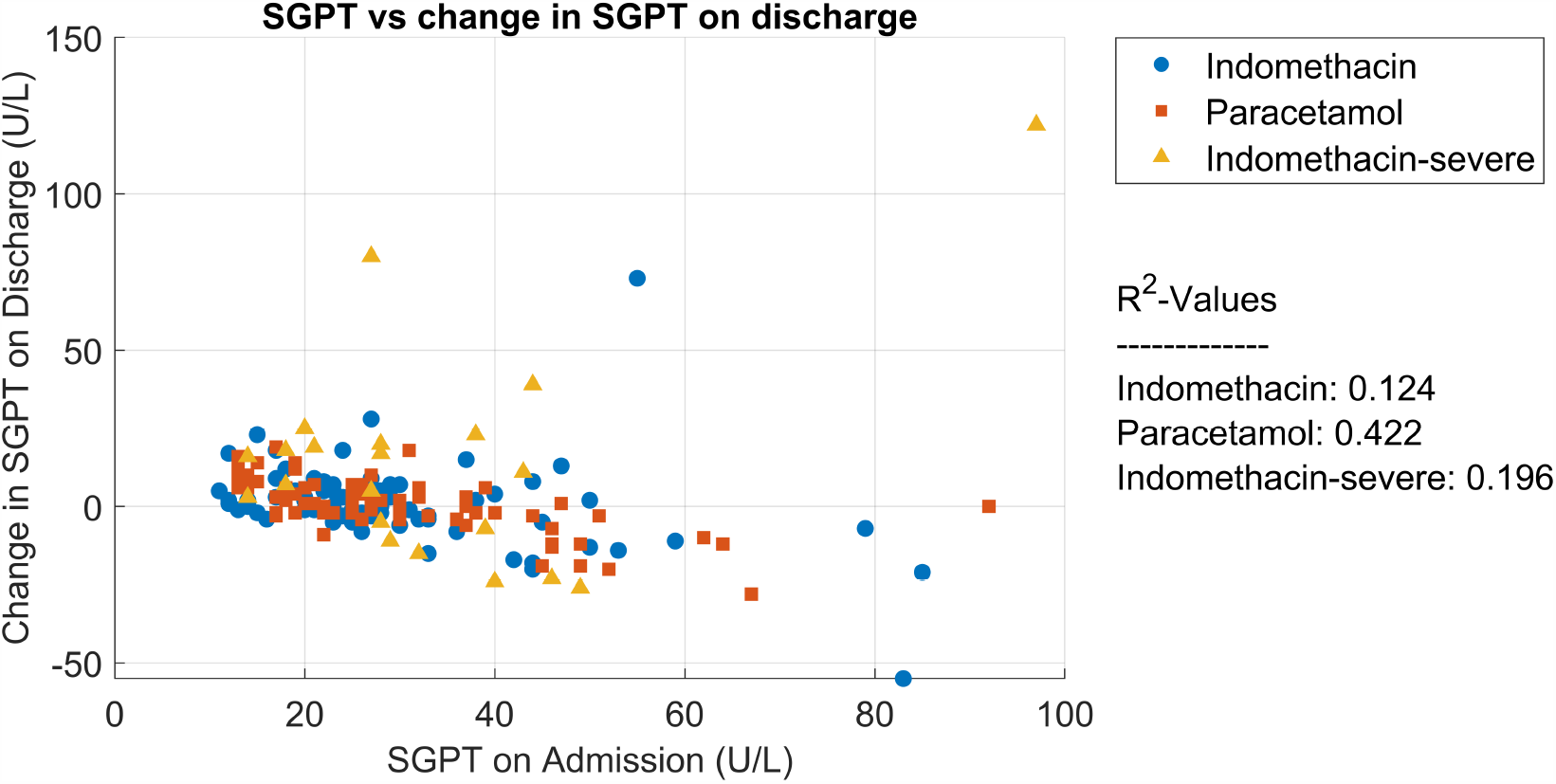
Change in SGPT after treatment as a function of SGPT on admission

All these figures show no deterioration after five days of Indomethacin except in one patient with CKD where the creatinine increased by 0.5 mg/dL. No other side-effects were reported by the patients or the attending physicians. Acute pancreatitis was seen in one patient in the severe group on admission. Indomethacin was continued for five days in that patient and he was discharged on the 17^th^ day after recovery. The prolonged stay was due to the pancreatitis rather than respiratory problem.

## 4.0 Discussion

The patients in the Indomethacin arm were found to have faster recovery compared to the paracetamol arm. As can be seen from Fig.8, many patients felt symptomatic relief with just two doses. Days for complete symptomatic recovery is three/four days in the former arm, compared to seven/eight days in the latter arm for symptomatic relief. Interestingly, even for severe cases, the symptomatic improvement is remarkable. The severe cases had received Indomethacin Sustained Release 75mg as against 25mg regular preparation twice a day in the mild and moderate cases. Age and CT-Score were observed to have no relation on the days for recovery.

The other objective of this study was to monitor the deterioration of patients to hypoxia. Of the eleven patients who complained of shortness of breath on admission in the Indomethacin arm only one was administered mild flow (2L/min) O_2_ for two days to give symptomatic relief. On the other hand, all the five patients in the paracetamol group who had shortness of breath on admission required supplementary oxygen. The results of the rest of the patients during the hospital stay are even more striking. A large proportion (34%), who did not have breathlessness on admission, but developed hypoxia requiring oxygen. Though they recovered, many had a prolonged stay in the hospital. None in the Indomethacin arm developed hypoxia. Supplementary oxygen was provided when Room Air (RA) SpO_2_ fell below 93% and the patient felt the need. Even more interesting is Fig. 13. The lone case which required oxygen in the first set was only after one day (second day after admission). Patients in the second arm, even after three-four days of treatment. deteriorated to hypoxia. Further analysis reported in Fig. 14 reveals that CT-Score and CRP at admission apart from shortness of breath at admission are factors in the second arm to develop hypoxia. Interaction between CRP at admission and CT score did not produce significant outcome. On the other hand, in the Indomethacin arm, with only one patient developing hypoxia CT-Score and other factors were overwhelmed by the treatment.

The hazard ratio for the indomethacin arm is 0.00218 with a 95% Confidence Interval of [0.00016, 0.031]. Lastly, prolonged stay in the hospital, beyond 14 days, is also a strong function of hypoxia at admission for the paracetamol arm. None in the Indomethacin group had a prolonged stay in the hospital. In the paracetamol arm twenty three patients had a prolonged stay. This conclusion is very important as the hospital infrastructure is not stretched by prolonged stay.

Reduction of C-Reactive Protein (CRP), a marker for inflammation, is plotted for patients in the Indomethacin arm. About 10% of the patients, who had low CRP levels on admission, had an elevated CRP after five days. Their follow-up after 14 days revealed no symptoms. This needs further investigation. Such a trend was not observed in the severe cases. Though the drug is five decades old, safety and adverse reactions were monitored through day-to-day observations and blood biochemistry. No patient developed nausea or vomiting, or gastro-intestinal bleeding in the form of malena. One patient admitted with nausea, vomiting and loose motions continued to have the symptoms during the treatment. She recovered after five days. She also developed mild hypoxia briefly but did not require supplementary oxygen.

There was no deterioration of renal or liver functions in the Indomethacin arm except in one patient with chronic kidney disease where the creatinine went up by 0.5% mg% from 1.2 mg%..

The study uses Propensity Score Matching, an accepted methodology in observational studies. Ethically, we felt that the study design is fair; it is close to RCT and blinded trials have been questioned for their inability to answer clinical questions [22]. Though the matched scores are acceptable on one metric, clinicians would like to view it on the actual covariates. Further insights are provided in Figs. 4 to 7. One can conclude from these figures that the patient profile distribution is similar in both the arms and the severe cases have higher CT-Scores and CRP.

The use of Indomethacin in 22 patients falling under the severe category was analysed separately, They received Indomethacin 75mg Sustained Release for five days along with Remedisvir for five days. Although these patients were hypoxemic on admission, they showed rapid symptomatic relief. However, they took longer time to recover from hypoxia and these patients could be discharged by 14 days except the patient with pancreatitis who stayed for 17 days. The cause of pancreatitis was not clear but Indomethacin was also given to him for 5 days. It is unlikely that Indomethacin caused the pancreatitis as the drug is under trial for treatment for acute pancreatitis [23]. Nevertheless, none of them required addition of steroids or transfer to the ICU or ventilator support.

The anti-inflammatory action of Indomethacin is well known [24]. SARS-Cov-2 produces severe disease not due to cytopathic action, but due to inflammation [25]. Indomethacin is a unique drug with anti-viral and anti-inflammatory actions. It is unfortunate that early publications cautioned the use of NSAIDs for Covid-19 [26].

Our study does not differentiate the anti-viral and anti-inflammatory actions of Indomethacin. But the patients in the Indomethacin arm of the study became rapidly asymptomatic, and had far less probability of developing hypoxia (Fig.13) compared to the paracetamol arm. The major drawback of the study is that Indomethacin was not used alone for Covid-19 treatment.

## 5.0 Conclusions

A registered clinical trial for assessing the efficacy and safety of Indomethacin for RT-PCR positive Covid-19 patients was carried out at two different centres. Propensity Scores were used to match with the paracetamol group. Symptomatic relief of Indomethacin patients was remarkable compared to the paracetamol group. Only one of the 72 patients required supplementary oxygen, while 28 out of 72 patients in the paracetamol group required oxygen supplement. The odds ratio of 0.022 sums up the statistical significance of the results. A set of 22 patients administered supplementary oxygen on admission did not deteriorate to warrant ventilation. Indomethacin should replace paracetamol in the treatment protocol for Covid-19 in the absence of specific contraindication

## Data Availability

The patient data if required can be obtained after ethics committee clearance of the hospital

## 6.0 Acknowledgement

The authors acknowledge the generous funding for this study by Mr. Kris Gopalakrishnan, Alumnus, Indian Institute of Technology Madras. But for him this study would not have been possible We also acknowledge the help of Dr. Meena for administrative help during the beginning of the trial and Mr. Krishna Teja during the preparation of the manuscript.

## References

1. Gordan D.E. et. al., A SARS-CoV-2 protein interaction map reveals targets for drug repurposing, Nature 2020 583 (July 16) 459–468

2. X. Wang and Y. Guang, COVID-19 drug repurposing: A review of computational screening methods, clinical trials, and protein interaction assays, Med Res Rev. 2020 1–24. https://doi.org/10.1002/med.21728

3. Andri Frediansyah et.al., Antivirals for COVID-19: A critical review, Clinical Epidemiology and global health 2020, https://doi.org/10.1016/j.cegh.2020.07.006

4. Carmen Gil, et.al COVID-19: Drug Targets and Potential TreatmentsJournal of Medicinal Chemistry 2020 63 (21) 12359–12386 DOI: 10.1021/acs.jmedchem.0c00606

5. Soy M, et.al., Cytokine storm in COVID-19: pathogenesis and overview of anti-inflammatory agents used in treatment, Clinical Rheumatology 2020 39 2085–2094

6. D.E. Gordan et.al. Comparative host-coronavirus protein interaction networks reveal pan-viral disease mechanisms 2020, 10.1126/Science.abe9403

7. Amici C, Di Caro A, Ciucci A, Chiappa L Castilleti, et al. Indomethacin has potent antiviral activity against SARS coronavirus, Antivir Ther 2006 11 1021–1030

8. Napolitano F et.al., Computational Drug Repositioning and Elucidation of Mechanism of Action of Compounds against SARS-CoV-2, 2020 https://arxiv.org/abs/2004.07697v2

9. Raghav L, et.al, Effect of some steroidal & non-steroidal anti-inflammatory drugs on purified goat brain cathepsin L, Indian Jl of Med. Res. 1993 98 188–92.

10. Bour A.M. et al., Interaction of indomethacin with cytokine production in whole blood. Potential mechanism for a brain-protective effect, Exp. Gerentol 2000 35 (8) 1017–1024

11. Russel B. et al., Associations between immune-suppressive and stimulating drugs and novel COVID-19—a systematic review of current evidence, ecancer 2020 14:1022 https://doi.org/10.3332/ecancer.2020.1022

12. First MR, Schroeder TJ, Hariharan S, Alexander JW, Weiskittel P. The effect of indomethacin on the febrile response following OKT3 therapy. Transplantation. 1992 53(1) 91–94

13. Gaughan WJ, Francos BB, Dunn SR, Francos GC, Burke JF. A retrospective analysis of the effect of indomethacin on adverse reactions to orthoclone OKT3 in the therapy of acute renal allograft rejection. Am J Kidney Dis. 1994 24(3) 486–90

14. Xu T, Gao X, Wu Z, Selinger DW, Zhou Z Indomethacin has a potent antiviral activity against SARS CoV-2 in vitro and canine coronavirus in vivo (preprint) April 2020 https://doi.org/10.1101/2020.04.01.017624

15. Ravichandran Rajan et.al., Low dose Indomethacin for Symptomatic Treatment of Covid-19, International Journal of Medical Reviews and Case Reports, Letters to the Editor (2020)

16. Kanakaraj A and Rajan Ravichandran, Low Dose Indomethacin in the Outpatient Treatment of COVID-19 in Kidney Transplant Recipients—A Case Series, Open Access Library Journal 2020 7, e6860.

17. Austin P.C. An Introduction to Propensity Score Methods for Reducing the Effects of Confounding in Observational Studies, Multivariate Behavioral Research, 46:399–424, 2011

18. WHO Solidarity Trial Consortium Repurposed Antiviral Drugs for Covid-19 - Interim WHO Solidarity Trial Results, NEJM, DOI: 10.1056/NEJMoa2023184

19. Momekov G and Momekova D, Ivermectin as a potential COVID-19 treatment from the pharmacokinetic point of view. medRxiv preprint doi:https://doi.org/10.1101/2020.04.11.20061804;

20. Tushar V.S. Sample Size Estimation in Clinical Trials, Perspect Clin Res. 2010 1(2) 67– 69.

21. Marinella M.A., Indomethacin and resveratrol as potential treatment adjuncts for SARS-CoV-2/COVID-19, to appear in The International Journal of Clinical Practice (online version: https://doi.org/10.1111/ijcp.13535)

22. Anand, R, et.al., ‘Fool’s gold? Why blinded trials are not always best’, BMJ (Clinical research ed.) 2020 368: 6228. https://doi.org/10.1136/bmj.l6228

23. A trial of Indomethacin in acute pancreatitis, ClinicalTrials.gov.Identifier NCT03547232

24. Donnelly P et.al, Indomethacin in rheumatoid arthritis: an evaluation of its anti-inflammatory and side effects. Br Med J. 1967 Jan 14; 1(5532): 69–75. doi:10.1136/bmj.1.5532.69

25. Tay M.Z. et al., The trinity of Covid-19: immunity, inflammation and intervention, Nat Rev Immunol 2020 20 (6) 363–374. doi:10.1038/s41577-020-0311-8.

26. Russel B, et.al., COVID-19 and treatment with NSAIDs and corticosteroids: should we be limiting their use in the clinical setting? ecancer 2020, 14:1023 https://doi.org/10.3332/ecancer.2020.1023

